# Repeated sleep-medication reporting patterns and cognitive change in HRS and SHARE

**DOI:** 10.64898/2026.11.06.26367411

**Authors:** Shupeng Wang, Dongdong Lu

## Abstract

**Background:** Self-reported sleep-medication use has been inconsistently linked to cognitive decline, largely because sleep disturbances, affective symptoms, and treatment transitions co-evolve over time. Moreover, whether repeated sleep-medication reports across consecutive survey waves follow a monotonic dose-response gradient with subsequent cognitive aging remains unclear. We evaluated baseline, two-wave, and three-wave longitudinal patterns of sleep-medication reporting in relation to cognitive trajectories across two large aging cohorts in the United States and Europe.

**Methods:** We analyzed population-based data from adults aged ≥50 years in the US Health and Retirement Study (HRS) and the Survey of Health, Ageing and Retirement in Europe (SHARE). First, we examined associations between baseline sleep-medication use and longitudinal changes in standardized global cognition and memory scores (Analysis 1: HRS n = 16,323; SHARE n = 41,453). Second, we conducted a prespecified two-wave landmark analysis classifying participants by reported sleep-medication use across two consecutive visits (HRS waves 11–12; SHARE waves 4–5) as 0, 1, or 2 use-positive visits, and by reporting sequence (00, 01, 10, and 11), with cognitive trajectories modeled from the second visit onward (Analysis 2: HRS n = 13,915; SHARE n = 29,214). Linear mixed-effects models with random intercepts and slopes adjusted for baseline sociodemographic, lifestyle, and clinical covariates and all covariate-by-time interactions. Third, we extended the landmark window to three consecutive visits (HRS waves 11–13, 2012–2016; SHARE waves 6–8, 2015–2020) to separate sustained from transient reporting, adjusting for interview mode and testing equivalence of sustained use to non-use (Analysis 3: HRS n = 12,310; SHARE n = 24,692).

**Results:** In baseline analyses, sleep-medication use was associated with modestly faster decline in global cognition (HRS: β = −0.0060 SD/year, 95% CI −0.0103 to −0.0017; SHARE: β = −0.0034 SD/year, 95% CI −0.0070 to 0.0002) and memory (HRS: β = −0.0062 SD/year, 95% CI −0.0115 to −0.0009; SHARE: β = −0.0037 SD/year, 95% CI −0.0074 to 0.0001). In the two-wave landmark analysis, reporting sleep-medication use at both visits (2 vs 0 visits) was not associated with accelerated cognitive decline in either cohort, refuting a monotonic consistency gradient. Instead, reporting use at a single visit (1 vs 0 visits) was associated with steeper decline in Western Europe (global cognition: β = −0.0087 SD/year, 95% CI −0.0157 to −0.0016; memory: β = −0.0083 SD/year, 95% CI −0.0158 to −0.0007) and among SHARE participants with secondary/vocational or tertiary education (interaction P = 0.041 for global cognition and P = 0.017 for memory). In the three-wave extension, no exposure contrast survived familywise correction; sustained three-visit reporting was statistically equivalent to persistent non-use within ±0.05 SD/year in both cohorts (equivalence P ≤ 0.0012).

**Conclusions:** Baseline sleep-medication reporting was associated with small negative differences in cognitive trajectories, whereas a fixed two-wave landmark design did not support a monotonic cumulative-exposure gradient. The localization of steeper cognitive decline to intermittent (one-visit) reporting in Western Europe and higher educational strata points to the role of treatment transitions and symptom instability rather than sustained medication exposure. Three-wave, mode-adjusted and equivalence-tested analyses indicate that sustained sleep-medication reporting is not accompanied by accelerated cognitive decline at the population level, whereas transient reporting continues to mark periods of elevated risk.

## Background

Cognitive decline and dementia impose a growing public-health burden as populations age: the global number of people living with dementia was estimated at 57.4 million in 2019 and is projected to reach 152.8 million by 2050 [1]. Insomnia disorder affects approximately one in five older adults [2], and the 2024 Lancet Commission on dementia prevention highlights sleep-related factors within the broader domain of modifiable risk [3]. Pharmacological sleep aids are widely used across aging populations: in the United States, 8.8% of adults aged 65 years and older report taking prescription sleep medications most days or every day [4], and in Europe benzodiazepine and Z-drug prescribing remains prevalent in both community-dwelling and long-term care settings [5,6].

Multiple biological pathways link sleep disturbance and cognitive aging, including impaired glymphatic clearance, β-amyloid and tau accumulation, and neuroinflammation. Concurrently, sedative-hypnotic medications may influence cognitive performance through central GABAergic modulation, acute sedation, or anticholinergic activity, while the underlying indications for treatment—insomnia, anxiety, and depressive symptoms—are themselves established correlates or prodromal features of cognitive decline. Because pharmacological exposure and underlying symptom severity co-evolve over time, longitudinal designs that distinguish stable, repeated reporting from treatment transitions offer greater analytical clarity than single cross-sectional assessments.

Observational studies of sleep medications and cognitive outcomes have yielded heterogeneous results. While several meta-analyses and registry-based investigations have linked benzodiazepine exposure—particularly long-term or high-cumulative-dose regimens—to elevated dementia risk [7–11], findings in nationally representative [12] and community-based cohorts [13–15] have been more variable, including null associations for cumulative dose among older adults with affective disorders or long-term follow-up [15,16]. In the Health and Retirement Study (HRS), sleep-treatment initiation has shown complex associations with subsequent cognitive trajectories that vary by baseline cognitive status [17]. Simultaneously, extreme sleep durations [18,19], anticholinergic medication burden [20,21], and longitudinal trajectories of sleep disturbance and multimorbidity [22,23] have each been implicated in late-life cognitive progression.

Across this literature, three methodological dimensions consistently shape observed associations: whether exposure is captured through cumulative dispensing records or single-wave self-reports, whether outcome follow-up is anchored strictly after exposure ascertainment, and how thoroughly underlying sleep and affective symptoms are controlled. Studies relying on long-term dispensing histories typically detect stronger associations [7,8], whereas single baseline self-reports in population surveys often yield smaller or null estimates [15,16]. Separating the timing of exposure ascertainment from subsequent cognitive follow-up is therefore essential to distinguish persistent exposure from acute treatment transitions and protopathic bias.

Sleep-medication use in older adults is also patterned by sex, depressive symptomatology, educational attainment, and regional prescribing practices. Women experience higher rates of both insomnia symptoms and hypnotic prescribing, and depressive symptoms correlate closely with both sleep complaints and cognitive change. Cross-national differences in healthcare access and deprescribing policies further shape who initiates, continues, or discontinues hypnotic therapy across US and European settings.

Three specific gaps remain unaddressed. First, most longitudinal cohort studies define exposure at a single baseline wave or aggregate reports across waves that overlap with cognitive follow-up, a practice susceptible to immortal-time and time-varying confounding. Anchoring cognitive follow-up at an explicit landmark visit after a fixed exposure window eliminates temporal overlap between exposure classification and outcome assessment. Second, single-wave exposure definitions conflate sustained use across waves with initiation or discontinuation, leaving unclear whether repeated sleep-medication reporting follows a monotonic dose-response gradient with subsequent cognitive change. Third, whether educational attainment and European regional prescribing contexts modify these longitudinal associations has rarely been evaluated within a harmonized cross-cohort framework. Recent multicohort investigations of functional and metabolic aging demonstrate the value of parallel HRS and SHARE analyses for testing cross-national generalizability [24,25].

Both HRS and SHARE are population-based longitudinal studies of adults aged 50 years and older with harmonized cognitive batteries, biennial interview schedules, and comprehensive sociodemographic and clinical assessments. HRS provides a nationally representative sample of US older adults, whereas SHARE comprises probability samples across participating European countries, enabling structured comparisons across European macro-regions.

Here, we integrated three prespecified analyses in the US Health and Retirement Study (HRS) and the Survey of Health, Ageing and Retirement in Europe (SHARE). Analysis 1 evaluated baseline sleep-medication reporting in relation to subsequent cognitive trajectories across the full follow-up period (HRS n = 16,323; SHARE n = 41,453). Analysis 2 applied a fixed two-wave exposure window (HRS waves 11–12; SHARE waves 4–5) with a landmark time zero at the second visit, classifying participants by both the count (0, 1, or 2 visits) and sequence (00, 01, 10, or 11) of reported use, and evaluated effect modification by educational attainment and European region (HRS n = 13,915; SHARE n = 29,214). Analysis 3 extended the landmark window to three consecutive biennial visits (HRS waves 11–13; SHARE waves 6–8) with follow-up anchored at the third visit, decomposing exposure into four cumulative counts (0– 3 visits), six clinical transition phenotypes, and all eight three-visit reporting sequences (000– 111), while adjusting for interview mode and formally testing statistical equivalence between sustained three-visit reporting and persistent non-use (HRS n = 12,310; SHARE n = 24,692). Across the landmark analyses, we tested two competing hypotheses: under a cumulative-exposure hypothesis, sustained multi-wave reporting (2 visits in Analysis 2; 3 visits [pattern 111] in Analysis 3) would predict the steepest cognitive decline along a monotonic gradient; conversely, under a treatment-transition and symptom-instability hypothesis, steeper cognitive decline would localize to transient or intermittent reporting rather than sustained multi-wave use.

## Methods

### Data sources and participants

We analyzed data from the RAND HRS Longitudinal File 2020 version 2, the Harmonized HRS version D, the Harmonized SHARE version F2, and SHARE release 9.0.0 [26–29]; cohort sampling, survey procedures, and data resource profiles have been detailed previously [30,31]. All cognitive outcomes, covariates, interview dates, and longitudinal medication indicators were reconstructed directly from the official primary cohort releases. For the baseline exposure analysis (Analysis 1), time zero was set at the first exposure visit (HRS wave 11, 2012; SHARE wave 4, 2011): participants aged ≥50 years with a valid baseline sleep-medication report, complete baseline covariates, and at least one valid cognitive assessment from baseline onward were included (HRS n = 16,323 for global cognition; SHARE n = 41,453). For the two-wave landmark analysis (Analysis 2), the exposure window spanned waves 11 (2012) and 12 (2014) in HRS and waves 4 (2011) and 5 (2013) in SHARE; participants aged ≥50 years at the first visit with valid sleep-medication reports at both visits, complete baseline covariates, a valid cognitive assessment at the second (landmark) visit, and at least one subsequent follow-up wave were included (HRS n = 13,915; SHARE n = 29,214; Figure S1; Table S1). For the three-wave landmark extension (Analysis 3), exposure was assessed across three consecutive biennial waves—waves 11–13 (2012–2016) in HRS and waves 6–8 (2015–2020) in SHARE—with cognitive follow-up anchored at the third visit (HRS wave 13; SHARE wave 8; HRS n = 12,310; SHARE n = 24,692).

### Study design and analytical framework

To compare conventional single-baseline exposure modeling with transition-resolved longitudinal frameworks, we prespecified three complementary analytical designs. Analysis 1 estimated the long-term association between baseline sleep-medication reporting and cognitive trajectories over all available waves from the first visit onward. Analysis 2 classified participants across two consecutive biennial visits and anchored cognitive follow-up strictly at the second visit (landmark time zero), ensuring that exposure ascertainment preceded outcome follow-up without temporal overlap and separating two-visit consistency from single-visit transitions (Figure 1a). Analysis 3 extended exposure classification across three consecutive visits and anchored cognitive follow-up at the third visit, distinguishing sustained three-visit use from late initiation, early discontinuation, mid-window single episodes, and on–off cycling.

**Figure 1.**
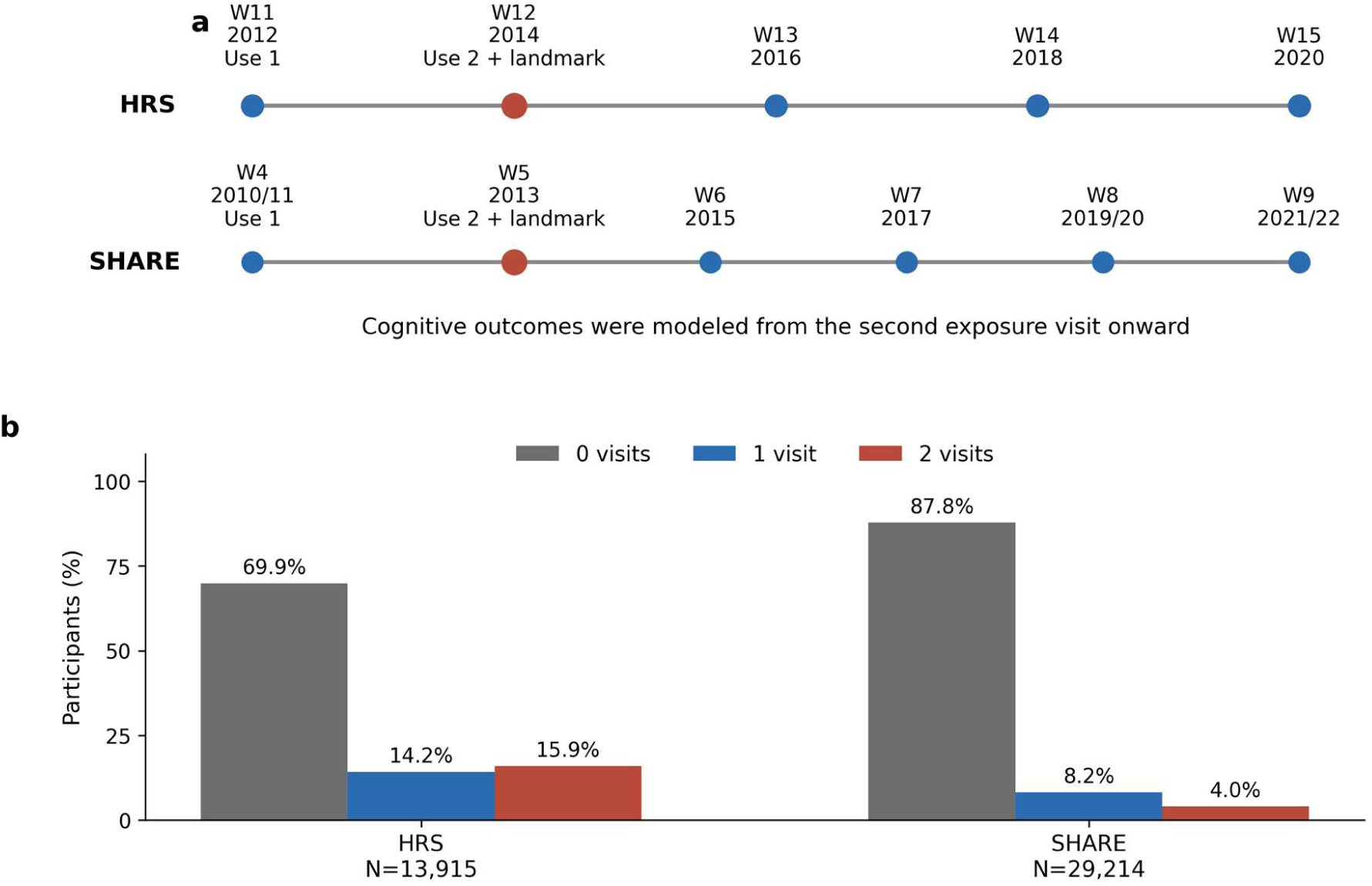
Two-wave landmark design and reporting distribution. Panel a illustrates the two consecutive exposure visits and subsequent cognitive follow-up waves in HRS and SHARE. Panel b displays the unweighted distribution of 0, 1, and 2 use-positive visits in the global-cognition landmark samples. The second exposure visit serves as the outcome landmark (time zero).

### Exposure ascertainment and landmark definition

In HRS, sleep-medication use was assessed in the core health module, where participants reported whether they currently take medication regularly to help with sleep. In SHARE, use was assessed via the standardized medication inventory (item PH011), where participants indicated whether they currently take drugs at least once a week for conditions presented on a showcard, including medication for sleep problems (category 9). Primary exposure was categorized by the number of visits within the two-wave window at which sleep-medication use was reported (0, 1, or 2 visits). In sequence-resolved analyses, exposure was further decomposed into four longitudinal patterns across visits 1 and 2: 00 (neither visit), 01 (visit 2 only), 10 (visit 1 only), and 11 (both visits). Time zero for all landmark trajectory models was defined as the second exposure visit.

In both cohorts, the two exposure assessments were separated by approximately two years. Because core survey modules in HRS and SHARE capture current self-reported regular or weekly use at each interview rather than continuous dispensing logs, drug classes, or daily dosages, the two-wave exposure classification characterizes longitudinal reporting consistency and visit-to-visit transitions across consecutive biennial assessments. Anchoring cognitive trajectories at the second exposure visit satisfies the temporal ordering required for longitudinal landmark analyses and precludes immortal-time bias.

### Cognitive outcomes and covariates

Primary outcomes were cohort-specific standardized global cognition and memory scores. In HRS, global cognition was operationalized using the validated 27-point composite score comprising immediate word recall (0–10), delayed word recall (0–10), Serial 7s subtraction (0– 5), and backward counting (0–2); memory was the sum of immediate and delayed word recall (0–20). In SHARE, global cognition combined immediate word recall (0–10), delayed word recall (0–10), and animal category fluency, while memory was the sum of immediate and delayed word recall (0–20). Within each cohort, raw cognitive scores across all waves were standardized to the mean and standard deviation of all eligible participants with a valid cognitive assessment at the landmark visit prior to requiring subsequent longitudinal follow-up; accordingly, regression slope coefficients represent annual change in standard deviation units (SD/year), and landmark baseline means in the retained longitudinal sample reflect modest positive selection (+0.1 SD in HRS; +0.2 SD in SHARE). Baseline covariates, ascertained at the first exposure visit, included age, sex, educational attainment (harmonized via ISCED-97 into lower than upper secondary, upper secondary or vocational, and tertiary), partnered status, body mass index (BMI), current smoking, alcohol consumption, depressive symptoms (8-item CES-D in HRS; 12-item EURO-D in SHARE), self-rated health (1 = excellent to 5 = poor), history of hypertension, diabetes, chronic lung disease, heart disease, stroke, and cancer, and concurrent use of other psychotropic medications (tranquilizers, antidepressants, or psychiatric drugs). All SHARE models additionally adjusted for country fixed effects.

### Statistical analysis

Following a prespecified statistical analysis plan (SAP) finalized prior to fitting the landmark models, we defined all exposure categories, covariate adjustments, effect-modification tests, minimum subgroup cell-size thresholds (n ≥ 100), and familywise Holm multiplicity corrections. Longitudinal cognitive trajectories were estimated using maximum-likelihood linear mixed-effects models with participant-specific random intercepts and random time slopes. Fixed effects included follow-up time (years since the landmark visit), exposure group, exposure-by-time interaction terms, all baseline covariates, and all covariate-by-time interactions; SHARE models additionally included country and country-by-time interactions. Primary parameters of interest were the exposure-by-time interaction coefficients, representing differences in annual cognitive change (SD/year) relative to the reference group (0 use-positive visits).

Effect modification by educational attainment was evaluated by incorporating full three-way interaction terms (exposure × time × education) and testing joint four-degree-of-freedom likelihood-ratio contrasts across the three educational strata. Regional modification in SHARE was evaluated within a pooled four-region interaction model (Western, Eastern, Southern, and Northern Europe, adjusted for country and country-by-time terms), with Western Europe prespecified as the primary regional focus and evaluated via leave-one-country-out replications. Sequence-resolved models estimated trajectories for patterns 01, 10, and 11 relative to 00, alongside direct pairwise contrasts among use-positive sequences (11 vs 01, 11 vs 10, and 01 vs 10). Holm step-down adjustments were applied within each prespecified hypothesis family. Because cognitive test batteries differ slightly between HRS and SHARE, cohort-specific models served as the primary analyses, complemented by inverse-variance fixed-effect pooled summaries. Reporting adheres to the Strengthening the Reporting of Observational Studies in Epidemiology (STROBE) cohort guideline [32] (Table S14).

All fitted models underwent formal diagnostics for random-effects singularity, convergence, and fixed-effect rank deficiency; all primary and sensitivity models converged with full rank (Table S9).

To evaluate robustness against sample selection, reverse causation, concurrent polypharmacy, sleep-related depressive symptoms, and non-specific chronic medication reporting, we conducted five prespecified sensitivity and supplementary analyses. First, we compared baseline characteristics between retained participants and those excluded due to missing post-landmark follow-up, estimated inverse-probability-of-inclusion weights (1/P[inclusion | baseline covariates], with predicted probabilities bounded at [0.02, 0.98] and weights winsorized at the 1st and 99th percentiles), and refit the landmark mixed-effects models reporting Kish effective sample sizes (Table S2a). Second, to test whether prior cognitive level or pre-landmark cognitive decline predicted subsequent sleep-medication reporting patterns, we fit multivariable multinomial logistic regression models with two-wave exposure count (0, 1, or 2 visits) as the outcome and visit-1 cognitive score and visit-1-to-landmark cognitive change (per 1 SD) as predictors (Table S2b). Third, we refit the landmark models omitting psychotropic co-medication (core model), adjusting for visit-1 baseline cognition and its time interaction, and controlling for the concurrent number of non-sleep, non-psychotropic medication classes (15 classes in HRS; 10 classes in SHARE) and its time interaction (Table S4). Fourth, in SHARE, where item-level EURO-D responses are harmonized across waves, we partitioned the 12-item scale into the dedicated sleep-trouble item (euro5) and the remaining 11-item non-sleep depressive score, entering both with time interactions (Table S4). Fifth, we repeated the exact two-wave landmark modeling framework using antihypertensive and lipid-lowering medications as negative-control chronic exposures (Table S4b). Detailed supplementary methods and complete model outputs are provided in Additional file 1.

### Three-wave landmark extension (Analysis 3)

To separate sustained exposure from transient reporting over a longer interval, we prespecified a three-wave extension in which exposure was ascertained across three consecutive biennial visits (HRS waves 11, 12, and 13, 2012–2016; SHARE waves 6, 7, and 8, 2015–2020), with the third visit (HRS wave 13; SHARE wave 8) serving as the landmark and cognitive trajectories modeled from that visit onward. Participants were classified by their three-visit reporting sequence (000, 001, 010, 011, 100, 101, 110, and 111), by the number of use-positive visits (0, 1, 2, or 3; reference = 0), and by six prespecified phenotypes: persistent non-use (000); late initiation (001/011); early discontinuation (100/110); mid-window single episode (010); on– off cycling (101); and sustained chronic use (111). The three-wave exposure samples comprised 12,594 HRS participants (35,144 cognitive observations; 68.0% reporting no use at any visit and 8.4% sustained use) and 24,692 SHARE participants (79,276 observations; 84.8% and 2.5%, respectively); after complete covariate ascertainment, 12,310 HRS and 24,692 SHARE participants entered the adjusted models (Table S10).

Models replicated the Analysis 1 and Analysis 2 specification, incorporating the same baseline covariates and covariate-by-time interactions, country and country-by-time terms in SHARE, and participant-specific random intercepts and random slopes, with follow-up anchored at the third exposure visit. Because both cohorts modified interview administration modes during the COVID-19 period, models additionally adjusted for face-to-face computer-assisted personal interviewing (CAPI) versus telephone or web-based interview mode and its time interaction, complemented by a sensitivity analysis restricted to face-to-face interviews (Table S12). Multiplicity was controlled by Holm step-down adjustment across the 12 primary three-wave count contrasts (two cohorts × two outcomes × three exposure levels) and, for the secondary families, within each cohort-outcome set (five phenotype contrasts in Table S10; seven sequence contrasts in Table S11). To evaluate whether sustained three-visit use (111) was statistically indistinguishable from persistent non-use (000), we performed two one-sided equivalence tests (TOST) against a prespecified primary equivalence margin of ±0.05 SD/year (and a secondary margin of ±0.10 SD/year), alongside one-sided non-inferiority tests (Table S13, Part 1).

Two additional analyses evaluated selection into the three-wave sample and reverse causation. First, inverse-probability weights for completing all three exposure waves were estimated from baseline sociodemographic and clinical covariates, with predicted probabilities bounded at [0.02, 0.98], weights winsorized at the 1st and 99th percentiles, and normalized to a mean of 1.0; weighted mixed-effects models were refit and Kish effective sample sizes reported (Table S12). Second, multivariable multinomial logistic regression models related pre-landmark global cognition and memory scores (scaled per 1 SD higher score) to subsequent three-wave exposure phenotypes (reference = persistent non-use), with the two cognitive domains modeled separately to avoid collinearity and adjusted for all baseline covariates (Table S13, Part 2).

## Results

### Participants and exposure distribution

The global-cognition landmark analytic samples comprised 13,915 participants in HRS (48,872 cognitive observations; mean 3.5 assessments per participant) and 29,214 participants in SHARE (97,115 observations; mean 3.3 assessments per participant); the corresponding memory samples included 13,891 and 29,821 participants, respectively (Figure S1; Table S1; Table S2). In HRS, 9,727 (69.9%), 1,970 (14.2%), and 2,218 (15.9%) participants reported sleep-medication use at 0, 1, and 2 visits, respectively; in SHARE, the corresponding counts were 25,638 (87.8%), 2,409 (8.2%), and 1,167 (4.0%) (Figure 1b). Sustained two-visit reporting was thus nearly fourfold more prevalent in the US cohort than in the European cohort. Across both cohorts, participants reporting sleep-medication use at one or two visits were older in SHARE, more frequently women, and had higher baseline depressive-symptom scores, poorer self-rated health, and substantially higher concurrent psychotropic medication use than non-users (Table 1).

**Table 1.** Baseline characteristics of the two-wave landmark samples by reported sleep-medication use *HRS (waves 11–12, 2012–2014)*

| **Characteristic** | **All**<br>**(n = 13,915)** | **0 visits**<br>**(n = 9,727)** | **1 visit**<br>**(n = 1,970)** | **2 visits**<br>**(n = 2,218)** | **P** |
|---|---|---|---|---|---|
| Age, mean (SD), years | 65.6 (9.7) | 65.6 (9.6) | 65.8 (10.0) | 65.5 (10.0) | 0.592 |
| Body mass index, mean (SD) | 28.9 (6.1) | 28.9 (6.0) | 29.2 (6.5) | 28.8 (6.5) | 0.054 |
| Depressive-symptom score, mean (SD) | 1.4 (2.0) | 1.1 (1.7) | 1.9 (2.2) | 2.5 (2.5) | <0.001 |
| Self-rated health score, mean (SD) | 2.8 (1.0) | 2.6 (1.0) | 3.0 (1.1) | 3.1 (1.1) | <0.001 |
| Global cognition at first exposure visit, mean (SD) | 0.1 (0.9) | 0.2 (0.9) | 0.1 (1.0) | 0.1 (0.9) | <0.001 |
| Global cognition at landmark, mean (SD) | 0.1 (1.0) | 0.2 (1.0) | 0.0 (1.0) | 0.1 (1.0) | <0.001 |
| Sex — Women | 8,268 (59.4) | 5,402 (55.5) | 1,287 (65.3) | 1,579 (71.2) | <0.001 |
| Partnered — Yes | 9,049 (65.0) | 6,450 (66.3) | 1,242 (63.0) | 1,357 (61.2) | <0.001 |
| Current smoking — Yes | 1,977 (14.2) | 1,299 (13.4) | 301 (15.3) | 377 (17.0) | <0.001 |
| Alcohol use — Yes | 7,984 (57.4) | 5,687 (58.5) | 1,067 (54.2) | 1,230 (55.5) | <0.001 |
| Hypertension — Yes | 7,998 (57.5) | 5,396 (55.5) | 1,230 (62.4) | 1,372 (61.9) | <0.001 |
| Diabetes — Yes | 3,049 (21.9) | 2,109 (21.7) | 449 (22.8) | 491 (22.1) | 0.533 |
| Lung disease — Yes | 1,145 (8.2) | 623 (6.4) | 218 (11.1) | 304 (13.7) | <0.001 |
| Heart disease — Yes | 2,808 (20.2) | 1,758 (18.1) | 456 (23.1) | 594 (26.8) | <0.001 |
| Stroke — Yes | 918 (6.6) | 560 (5.8) | 155 (7.9) | 203 (9.2) | <0.001 |
| Cancer — Yes | 1,760 (12.6) | 1,162 (11.9) | 251 (12.7) | 347 (15.6) | <0.001 |
| Other psychotropic-medication use — Yes | 2,499 (18.0) | 948 (9.7) | 533 (27.1) | 1,018 (45.9) | <0.001 |
| Education — Lower than upper secondary | 2,299 (16.5) | 1,613 (16.6) | 346 (17.6) | 340 (15.3) | 0.195 |
| Upper secondary or vocational | 8,233 (59.2) | 5,723 (58.8) | 1,175 (59.6) | 1,335 (60.2) |  |
| Tertiary | 3,383 (24.3) | 2,391 (24.6) | 449 (22.8) | 543 (24.5) |  |
| Age, mean (SD), years | 64.9 (9.1) | 64.5 (8.9) | 67.6 (9.8) | 67.9 (10.0) | <0.001 |
| Body mass index, mean (SD) | 27.0 (4.7) | 26.9 (4.6) | 27.5 (5.5) | 27.1 (5.2) | <0.001 |
| Depressive-symptom score, mean (SD) | 2.4 (2.1) | 2.1 (2.0) | 3.8 (2.4) | 4.6 (2.5) | <0.001 |
| Self-rated health score, mean (SD) | 3.1 (1.1) | 3.0 (1.0) | 3.7 (1.0) | 3.9 (0.9) | <0.001 |
| Global cognition at first exposure visit, mean (SD) | 0.2 (0.9) | 0.2 (0.9) | −0.1 (1.0) | −0.1 (1.0) | <0.001 |
| Global cognition at landmark, mean (SD) | 0.2 (1.0) | 0.3 (1.0) | −0.1 (1.0) | −0.1 (1.0) | <0.001 |
| Sex — Women | 16,802 (57.5) | 14,188 (55.3) | 1,712 (71.1) | 902 (77.3) | <0.001 |
| Partnered — Yes | 20,380 (69.8) | 18,217 (71.1) | 1,495 (62.1) | 668 (57.2) | <0.001 |
| Current smoking — Yes | 5,293 (18.1) | 4,676 (18.2) | 379 (15.7) | 238 (20.4) | 0.001 |
| Alcohol use — Yes | 18,048 (61.8) | 16,244 (63.4) | 1,206 (50.1) | 598 (51.2) | <0.001 |
| Hypertension — Yes | 12,348 (42.3) | 10,429 (40.7) | 1,290 (53.5) | 629 (53.9) | <0.001 |
| Diabetes — Yes | 3,492 (12.0) | 2,859 (11.2) | 424 (17.6) | 209 (17.9) | <0.001 |
| Lung disease — Yes | 2,105 (7.2) | 1,653 (6.4) | 286 (11.9) | 166 (14.2) | <0.001 |
| Heart disease — Yes | 4,216 (14.4) | 3,343 (13.0) | 567 (23.5) | 306 (26.2) | <0.001 |
| Stroke — Yes | 1,248 (4.3) | 941 (3.7) | 195 (8.1) | 112 (9.6) | <0.001 |
| Cancer — Yes | 2,062 (7.1) | 1,671 (6.5) | 260 (10.8) | 131 (11.2) | <0.001 |
| Other psychotropic-medication use — Yes | 1,683 (5.8) | 892 (3.5) | 419 (17.4) | 372 (31.9) | <0.001 |
| Education — Lower than upper secondary | 11,056 (37.8) | 9,307 (36.3) | 1,131 (46.9) | 618 (53.0) | <0.001 |
| Upper secondary or vocational | 11,500 (39.4) | 10,288 (40.1) | 869 (36.1) | 343 (29.4) |  |
| Tertiary | 6,658 (22.8) | 6,043 (23.6) | 409 (17.0) | 206 (17.7) |  |

*Values are mean (SD) or n (%). The Table 1 sample is the outcome-complete global-cognition landmark sample. P values are descriptive one-way analysis-of-variance (ANOVA) or Pearson chi-square comparisons across the three reporting groups. Sleep-medication use was ascertained at HRS waves 11 (2012) and 12 (2014) and SHARE waves 4 (2011) and 5 (2013). Depressive symptoms were measured with the 8-item CES-D (0–8) in HRS and the 12-item EURO-D (0–12) in SHARE; self-rated health was rated from 1 (excellent) to 5 (poor) in both cohorts. Participants contributed an average of 3.5 (HRS) and 3.3 (SHARE) cognitive assessments from the landmark visit onward. Because cognitive scores were standardized within each cohort against all eligible participants with a valid landmark-visit assessment prior to requiring longitudinal follow-up, baseline landmark means (+0.1 SD in HRS; +0.2 SD in SHARE) reflect modest positive selection among participants with subsequent follow-up assessments.*

### Baseline sleep-medication use and cognitive trajectories (Analysis 1)

In single-baseline longitudinal models (Analysis 1), baseline sleep-medication reporting was associated with modestly faster annual decline in global cognition (HRS: β = −0.0060 SD/year, 95% CI −0.0103 to −0.0017, P = 0.007; SHARE: β = −0.0034 SD/year, 95% CI −0.0070 to 0.0002, P = 0.062) and memory (HRS: β = −0.0062 SD/year, 95% CI −0.0115 to −0.0009, P = 0.022; SHARE: β = −0.0037 SD/year, 95% CI −0.0074 to 0.0001, P = 0.053) (Table 2). Inverse-variance pooled estimates across the two cohorts were −0.0044 SD/year (95% CI −0.0072 to −0.0017; P = 0.0016) for global cognition and −0.0045 SD/year (95% CI −0.0076 to −0.0015; P = 0.0038) for memory. Baseline analyses in SHARE also indicated steeper memory decline in Western Europe (β = −0.0087 SD/year) and significant effect modification by educational attainment for memory (nominal P = 0.013; Holm-adjusted P = 0.027).

**Table 2.** Integrated baseline-use (Analysis 1), two-wave (Analysis 2) and three-wave (Analysis 3) landmark estimates.

| Analysis | Cohort | Outcome | N | Comparison | SD/year (95% CI) | P | Holm P |
|---|---|---|---|---|---|---|---|
| Analysis 1: Baseline exposure | HRS | Global cognition | 16,323 | Baseline use vs non-use | −0.0060 (−0.0103 to −0.0017) | 0.007 | — |
| Analysis 1: Baseline exposure | HRS | Memory | 16,314 | Baseline use vs non-use | −0.0062 (−0.0115 to −0.0009) | 0.022 | — |
| Analysis 1: Baseline exposure | SHARE | Global cognition | 41,453 | Baseline use vs non-use | −0.0034 (−0.0070 to 0.0002) | 0.062 | — |
| Analysis 1: Baseline exposure | SHARE | Memory | 42,311 | Baseline use vs non-use | −0.0037 (−0.0074 to 0.0001) | 0.053 | — |
| Analysis 2: Two-wave landmark | HRS | Global cognition | 13,915 | 1 vs 0 reported-use visits | −0.0013 (−0.0088 to 0.0063) | 0.744 | 1.000 |
| Analysis 2: Two-wave landmark | HRS | Global cognition | 13,915 | 2 vs 0 reported-use visits | −0.0036 (−0.0112 to 0.0040) | 0.356 | 1.000 |
| Analysis 2: Two-wave landmark | HRS | Memory | 13,891 | 1 vs 0 reported-use visits | 0.0016 (−0.0073 to 0.0105) | 0.725 | 1.000 |
| Analysis 2: Two-wave landmark | HRS | Memory | 13,891 | 2 vs 0 reported-use visits | −0.0073 (−0.0163 to 0.0017) | 0.112 | 0.679 |
| Analysis 2: Two-wave landmark | SHARE | Global cognition | 29,214 | 1 vs 0 reported-use visits | −0.0045 (−0.0092 to 0.0003) | 0.065 | 0.523 |
| Analysis 2: Two-wave landmark | SHARE | Global cognition | 29,214 | 2 vs 0 reported-use visits | 0.0016 (−0.0053 to 0.0085) | 0.656 | 1.000 |
| Analysis 2: Two-wave landmark | SHARE | Memory | 29,821 | 1 vs 0 reported-use visits | −0.0043 (−0.0094 to 0.0008) | 0.097 | 0.679 |
| Analysis 2: Two-wave landmark | SHARE | Memory | 29,821 | 2 vs 0 reported-use visits | 0.0015 (−0.0059 to 0.0090) | 0.691 | 1.000 |
| Analysis 3: Three-wave landmark | HRS | Global cognition | 12,310 | 1 vs 0 reported-use visits | −0.0134 (−0.0258 to −0.0010) | 0.034 | 0.410 |
| Analysis 3: Three-wave landmark | HRS | Global cognition | 12,310 | 2 vs 0 reported-use visits | −0.0043 (−0.0189 to 0.0103) | 0.564 | 1.000 |
| Analysis 3: Three-wave landmark | HRS | Global cognition | 12,310 | 3 vs 0 reported-use visits | −0.0147 (−0.0308 to 0.0014) | 0.074 | 0.740 |
| Analysis 3: Three-wave landmark | HRS | Memory | 12,310 | 1 vs 0 reported-use visits | −0.0129 (−0.0280 to 0.0022) | 0.093 | 0.836 |
| Analysis 3: Three-wave landmark | HRS | Memory | 12,310 | 2 vs 0 reported-use visits | −0.0116 (−0.0294 to 0.0062) | 0.202 | 1.000 |
| Analysis 3: Three-wave landmark | HRS | Memory | 12,310 | 3 vs 0 reported-use visits | −0.0186 (−0.0381 to 0.0009) | 0.061 | 0.672 |
| Analysis 3: Three-wave landmark | SHARE | Global cognition | 24,692 | 1 vs 0 reported-use visits | −0.0020 (−0.0092 to 0.0052) | 0.590 | 1.000 |
| Analysis 3: Three-wave landmark | SHARE | Global cognition | 24,692 | 2 vs 0 reported-use visits | −0.0065 (−0.0170 to 0.0040) | 0.226 | 1.000 |
| Analysis 3: Three-wave landmark | SHARE | Global cognition | 24,692 | 3 vs 0 reported-use visits | 0.0012 (−0.0121 to 0.0145) | 0.859 | 1.000 |
| Analysis 3: Three-wave landmark | SHARE | Memory | 24,692 | 1 vs 0 reported-use visits | −0.0008 (−0.0087 to 0.0072) | 0.851 | 1.000 |
| Analysis 3: Three-wave landmark | SHARE | Memory | 24,692 | 2 vs 0 reported-use visits | −0.0054 (−0.0170 to 0.0061) | 0.357 | 1.000 |
| Analysis 3: Three-wave landmark | SHARE | Memory | 24,692 | 3 vs 0 reported-use visits | −0.0006 (−0.0153 to 0.0140) | 0.933 | 1.000 |

*Analysis 1 (baseline exposure) models longitudinal cognitive trajectories from the first exposure visit onward; Analysis 2 (two-wave landmark) classifies exposure across two consecutive visits and models cognitive trajectories from the second (landmark) visit onward. Holm-adjusted P values in Analysis 2 account for multiplicity across the eight primary count-based slope contrasts. Negative coefficients (SD/year) indicate steeper annual cognitive decline relative to 0 reported-use visits. Analysis 3 (three-wave landmark) classifies exposure across three consecutive visits (HRS waves 11–13; SHARE waves 6–8) and models trajectories from the third visit onward; its Holm-adjusted P values derive from the 12-test primary three-wave count family.*

### Two-wave reporting patterns and subsequent cognitive change (Analysis 2)

In the two-wave landmark analysis, reporting sleep-medication use at both consecutive visits (2 vs 0 visits) was not associated with accelerated decline in either cohort (Table 2; Figure 2; Figure 3). In HRS, two-visit versus zero-visit slope differences were −0.0036 SD/year (95% CI −0.0112 to 0.0040; P = 0.356) for global cognition and −0.0073 SD/year (95% CI −0.0163 to 0.0017; P = 0.112) for memory; in SHARE, the corresponding estimates were +0.0016 SD/year (95% CI −0.0053 to 0.0085; P = 0.656) and +0.0015 SD/year (95% CI −0.0059 to 0.0090; P = 0.691). These findings did not support a monotonic 0-, 1-, and 2-visit cumulative exposure gradient.

**Figure 2.**
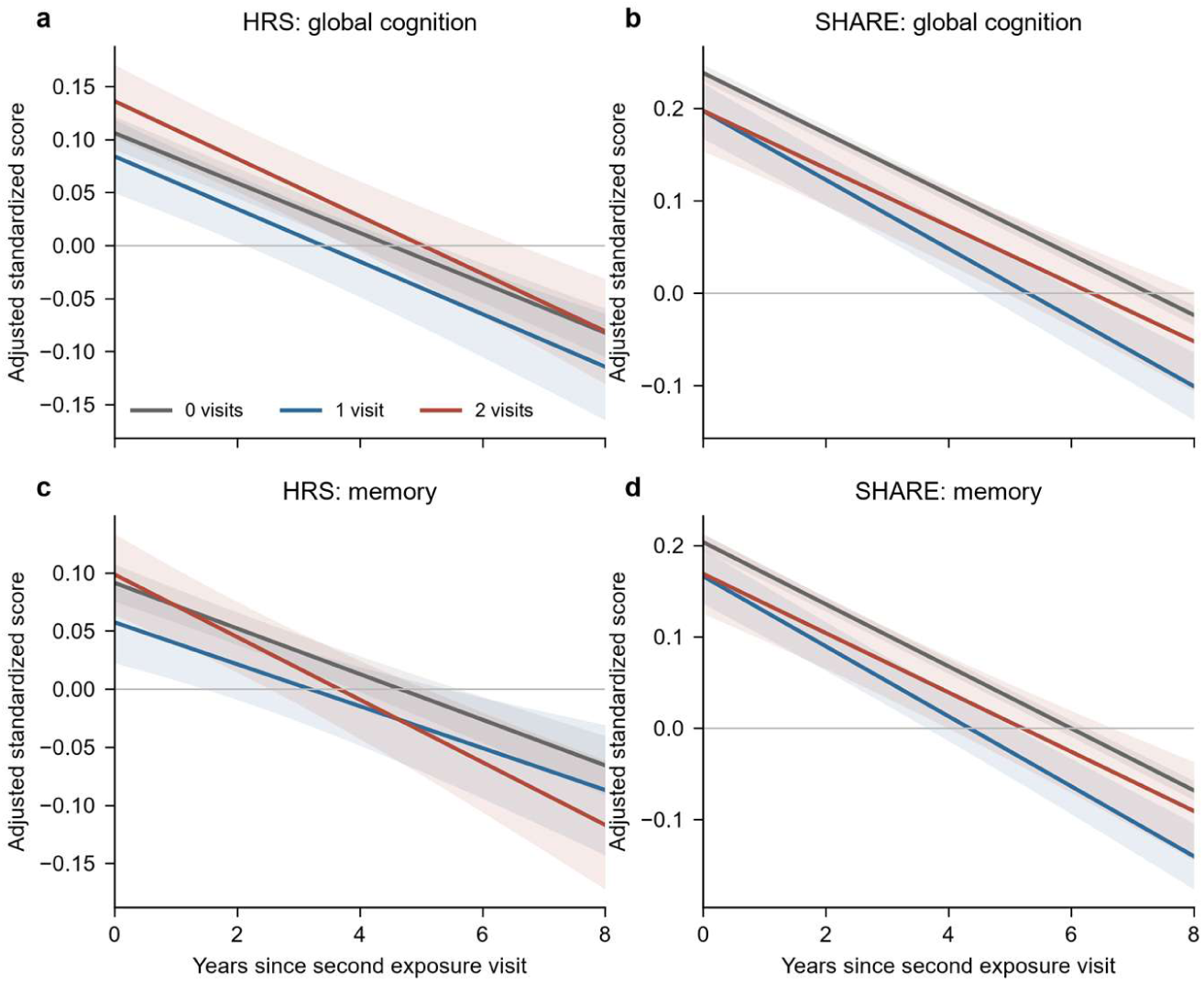
Adjusted cognitive trajectories by two-wave reporting group. Fixed-effect predicted cognitive trajectories evaluated at centered covariates from the fully adjusted covariate-by-time linear mixed-effects models. Shaded bands represent pointwise 95% confidence intervals. Intercepts at time zero reflect adjusted group differences at the landmark visit; slope contrasts are given in Table 2.

**Figure 3.**
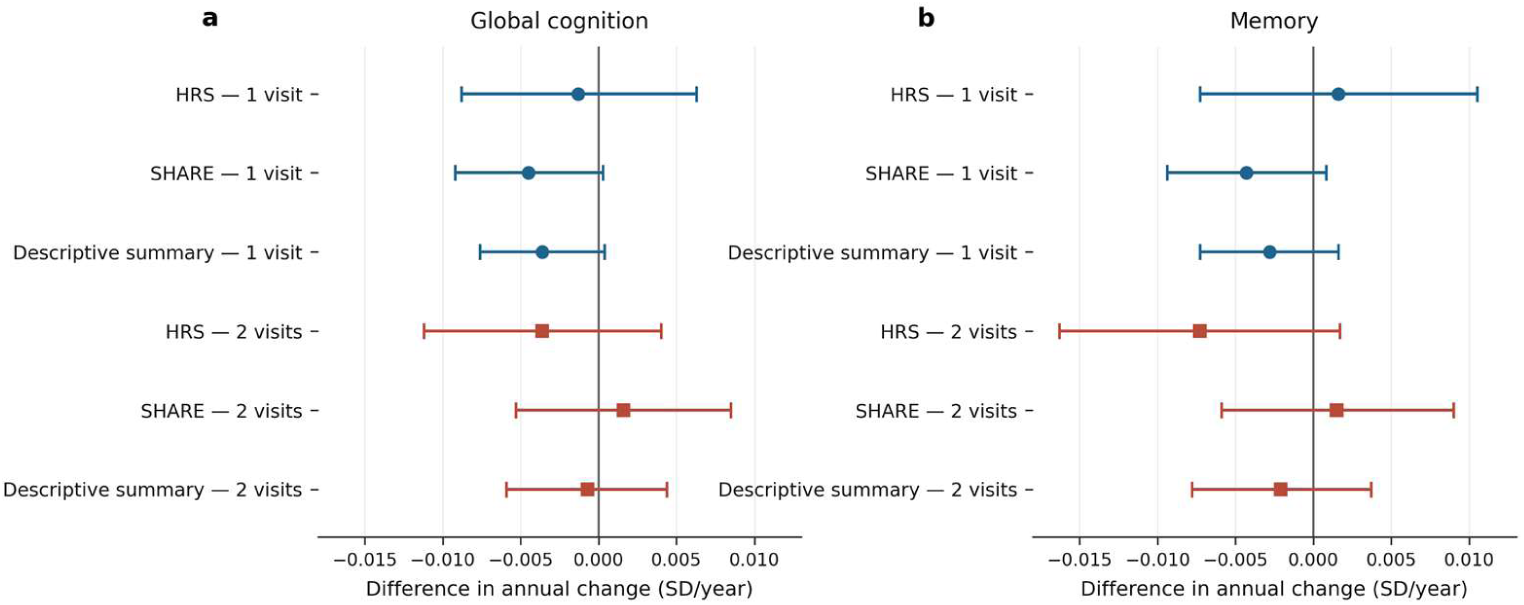
Overall two-wave reporting associations. Adjusted annual slope differences (SD/year) and 95% confidence intervals for 1 and 2 reported-use visits relative to 0 visits. Descriptive summaries combine cohort-specific HRS and SHARE slope differences via inverse-variance fixed-effects pooling.

For intermittent single-visit reporting (1 vs 0 visits), SHARE participants exhibited negative point estimates for both global cognition (β = −0.0045 SD/year, 95% CI −0.0092 to 0.0003; P = 0.065) and memory (β = −0.0043 SD/year, 95% CI −0.0094 to 0.0008; P = 0.097), whereas HRS single-visit estimates were close to zero (global cognition: β = −0.0013 SD/year, P = 0.744; memory: β = +0.0016 SD/year, P = 0.725). Across all eight primary cohort-by-outcome count contrasts, 95% confidence intervals overlapped zero and familywise Holm-adjusted P values exceeded 0.05 (Table 2). In sequence-resolved analyses (Table S3), both transition patterns in SHARE showed comparable negative point estimates relative to 00—for visit-2 initiation (01 vs 00), β = −0.0051 SD/year in global cognition and −0.0032 SD/year in memory; for visit-2 discontinuation (10 vs 00), β = −0.0038 and −0.0055 SD/year, respectively—whereas consistent two-visit reporting (11 vs 00) remained near zero (+0.0016 and +0.0015 SD/year), with no significant difference between patterns 01 and 10 (all P ≥ 0.237). In HRS memory, pattern 11 showed a nominally steeper slope than pattern 10 (β = −0.0142 SD/year, nominal P = 0.041; Holm P = 0.986 across the 24-test sequence family).

### Three-wave reporting patterns and subsequent cognitive change (Analysis 3)

In the three-wave extension, 12,594 HRS participants (61.7% of the 20,405 eligible at visit 1) and 24,692 SHARE participants (84.2% of the 29,306 eligible) contributed 35,144 and 79,276 cognitive observations (mean 2.8 and 3.2 per participant). Persistent non-use predominated (HRS 8,567 [68.0%]; SHARE 20,943 [84.8%]), and sustained three-visit use remained more common in HRS (1,057 [8.4%]) than in SHARE (621 [2.5%]); mid-window single episodes (010) occurred in 721 and 663 participants, and on–off cycling (101) in 121 and 275, respectively. Complete covariate data were available for 12,310 HRS and 24,692 SHARE participants, who form the analytic samples for all three-wave models.

Three-visit frequency contrasts are presented in Table 2. In HRS, each additional use-positive visit was associated with small negative global-cognition slope differences (1 vs 0: β = −0.0134 SD/year, 95% CI −0.0258 to −0.0010, P = 0.034; 2 vs 0: −0.0043 SD/year, 95% CI −0.0189 to 0.0103, P = 0.564; 3 vs 0: −0.0147 SD/year, 95% CI −0.0308 to 0.0014, P = 0.074), with corresponding memory estimates of −0.0129 SD/year, 95% CI −0.0280 to 0.0022 (P = 0.093), −0.0116 SD/year, 95% CI −0.0294 to 0.0062 (P = 0.202) and −0.0186 SD/year, 95% CI −0.0381 to 0.0009 (P = 0.061). None of the 12 primary count contrasts survived familywise correction (all Holm P ≥ 0.410). In SHARE, all count contrasts were close to zero (global cognition: −0.0020, −0.0065 and +0.0012 SD/year; memory: −0.0008, −0.0054 and −0.0006 SD/year; all P ≥ 0.23; all Holm P = 1.000).

In phenotype models (Table S10), the only nominally significant HRS contrasts were mid-window single episodes for global cognition (β = −0.0204 SD/year, 95% CI −0.0385 to −0.0023, P = 0.027) and memory (−0.0235 SD/year, 95% CI −0.0456 to −0.0013, P = 0.038); sustained chronic use showed smaller negative estimates that did not reach nominal significance (global −0.0146 SD/year, 95% CI −0.0307 to 0.0015, P = 0.076; memory −0.0183 SD/year, 95% CI −0.0378 to 0.0012, P = 0.066), and on–off cycling was null (+0.0175 SD/year, P = 0.435). No phenotype contrast reached familywise significance (smallest Holm P = 0.137). In SHARE, every phenotype estimate fell within ±0.008 SD/year and all nominal P values were ≥ 0.34.

In sequence-resolved models (Table S11), the HRS mid-window single episode (010) carried the largest negative deviation (global −0.0204 SD/year, 95% CI −0.0386 to −0.0023, P = 0.027; memory −0.0235 SD/year, 95% CI −0.0456 to −0.0014, P = 0.037), whereas sustained use (111) was intermediate (global −0.0147 SD/year, 95% CI −0.0308 to 0.0014, P = 0.074; memory −0.0184 SD/year, 95% CI −0.0378 to 0.0011, P = 0.065); no sequence contrast attained familywise significance (smallest Holm P = 0.190). SHARE sequence estimates were uniformly null (all nominal P ≥ 0.16; all Holm P = 1.000). Equivalence testing of sustained use versus persistent non-use (Table S13, Part 1) showed that the 95% confidence interval excluded the prespecified ±0.05 SD/year margin in both cohorts: HRS global −0.0149 SD/year, 95% CI −0.0312 to 0.0014 (equivalence P = 1.2 × 10⁻⁵) and memory −0.0192 SD/year, 95% CI −0.0390 to 0.0006 (P = 0.001); SHARE global 0.0021 SD/year, 95% CI −0.0112 to 0.0154 (P = 7.9 × 10⁻¹³) and memory 0.0006 SD/year, 95% CI −0.0141 to 0.0152 (P = 1.9 × 10⁻¹¹).

In sensitivity analyses (Table S12), inverse-probability weights for completing all three exposure waves had mean predicted probabilities of 0.80 in HRS and 0.46 in SHARE, with Kish effective sample sizes of 12,316 and 24,018 and maximum weights of 1.39 and 1.52; weighted slope estimates were virtually unchanged (HRS global cognition 1 vs 0 visits: β = −0.0134 SD/year, 95% CI −0.0258 to −0.0011, P = 0.033; HRS memory 3 vs 0 visits: β = −0.0194 SD/year, 95% CI −0.0385 to −0.0003, P = 0.046; all SHARE contrasts P ≥ 0.266). Restricting to face-to-face (CAPI) interviews left all SHARE estimates null (n = 12,079; all P ≥ 0.489); in the much smaller HRS face-to-face-only subset (n = 790 across all exposure groups), memory contrasts were null (all P ≥ 0.695) and global-cognition models were not estimated because exposed subgroup cells fell below the prespecified minimum threshold (n < 100) required for mixed-effects convergence. In multinomial reverse-causation models (Table S13, Part 2), higher pre-landmark cognitive scores in HRS were associated with significantly lower relative risk of early discontinuation (global cognition: RRR = 0.69 per 1 SD higher score, 95% CI 0.59–0.80, P < 0.001; memory: RRR = 0.81, 95% CI 0.71–0.92, P = 0.001) and mid-window single episodes (global cognition: RRR = 0.72, 95% CI 0.55–0.94, P = 0.014; memory: RRR = 0.79, 95% CI 0.63–0.99, P = 0.039)—indicating that lower pre-landmark cognition predicted transient use and discontinuation—whereas pre-landmark cognition was unrelated to sustained three-visit use (global cognition: RRR = 0.96, 95% CI 0.88–1.04, P = 0.293; memory: RRR = 1.04, 95% CI 0.98–1.11, P = 0.230). In SHARE, higher pre-landmark cognition was modestly associated with sustained three-visit reporting (global cognition: RRR = 1.22 per 1 SD higher score, 95% CI 1.01–1.48, P = 0.044; memory: RRR = 1.21, 95% CI 1.06–1.38, P = 0.004), while all transient phenotypes were null (all P ≥ 0.167).

### Regional concentration in Western Europe and modification by education

In the pooled SHARE regional interaction model, single-visit reporting (1 vs 0 visits) was associated with steeper decline in Western Europe for both global cognition (β = −0.0087 SD/year, 95% CI −0.0157 to −0.0016; P = 0.016) and memory (β = −0.0083 SD/year, 95% CI −0.0158 to −0.0007; P = 0.032) (Figure 4; Table S7). Leave-one-country-out analyses across Western European countries yielded consistent negative one-visit slope estimates ranging from −0.0098 to −0.0062 SD/year for global cognition and from −0.0091 to −0.0058 SD/year for memory, confirming that no single country drove the regional signal (Table S8). Omnibus regional interaction tests across all four European macro-regions did not reach statistical significance (global cognition P = 0.212; memory P = 0.114; Holm P = 0.229 for both; Table S6).

**Figure 4.**
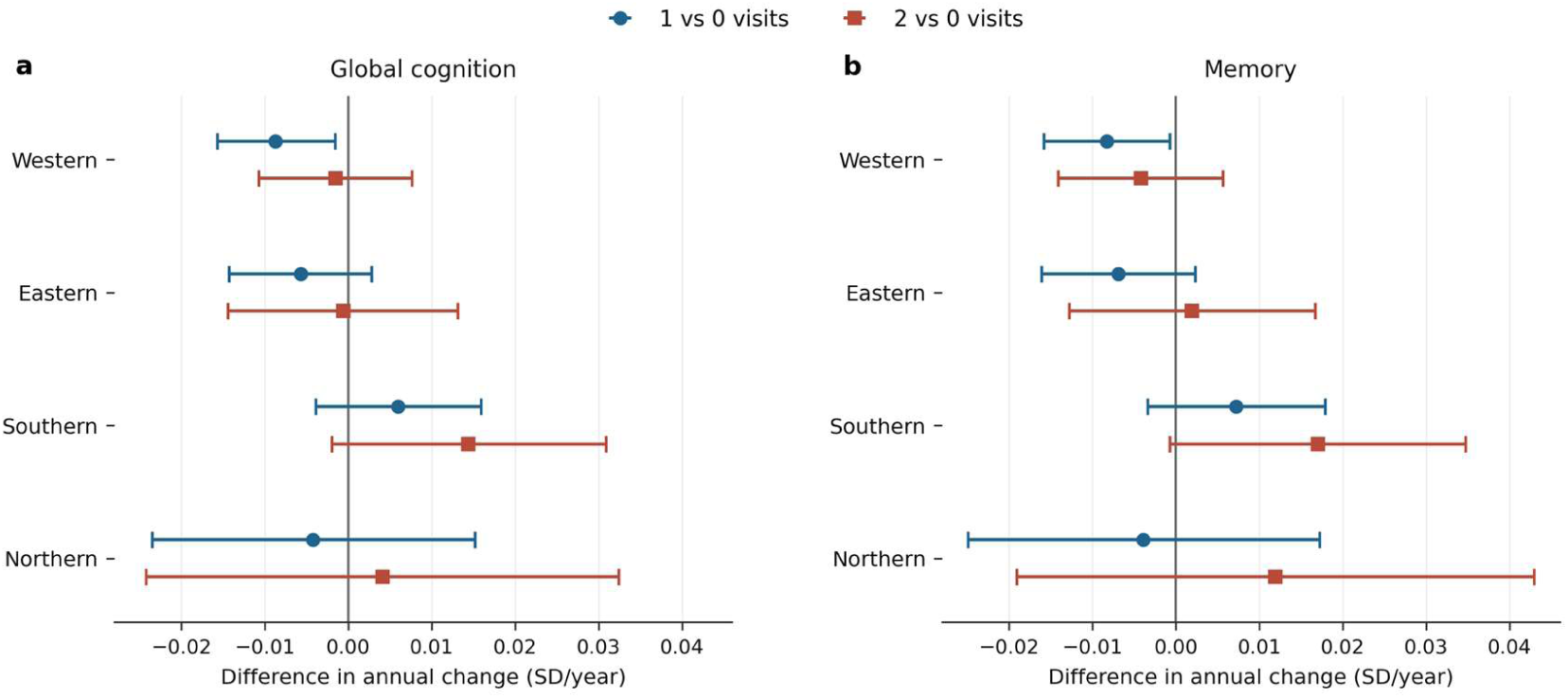
Two-wave reporting associations across SHARE regions. Adjusted annual slope differences (SD/year) and 95% confidence intervals across all four European macro-regions from the pooled exposure-by-region-by-time interaction model adjusted for country and country-by-time terms. One-visit confidence intervals in Western Europe exclude zero for both global cognition and memory; omnibus regional interaction P values are reported in Table S6.

In SHARE, educational attainment modified the two-wave association at nominal significance for both global cognition (joint 4-df interaction P = 0.041; Holm P = 0.124) and memory (joint P = 0.017; Holm P = 0.067) (Figure 5; Table S6). Specifically, for 1 versus 0 reported-use visits in SHARE, global-cognition slope differences were +0.0032 SD/year (P = 0.370) in lower education, −0.0108 SD/year (95% CI −0.0183 to −0.0033; P = 0.005) in secondary/vocational education, and −0.0094 SD/year (95% CI −0.0203 to 0.0014; P = 0.088) in tertiary education; the corresponding memory estimates were +0.0049 SD/year (P = 0.204), −0.0101 SD/year (95% CI −0.0182 to −0.0020; P = 0.014), and −0.0145 SD/year (95% CI −0.0262 to −0.0028; P = 0.015), respectively (Table S5). In HRS, educational interaction tests were null (P = 0.735 for global cognition; P = 0.436 for memory), although a nominal two-visit memory association was observed within the secondary/vocational stratum (β = −0.0121 SD/year, 95% CI −0.0233 to −0.0009; P = 0.034).

**Figure 5.**
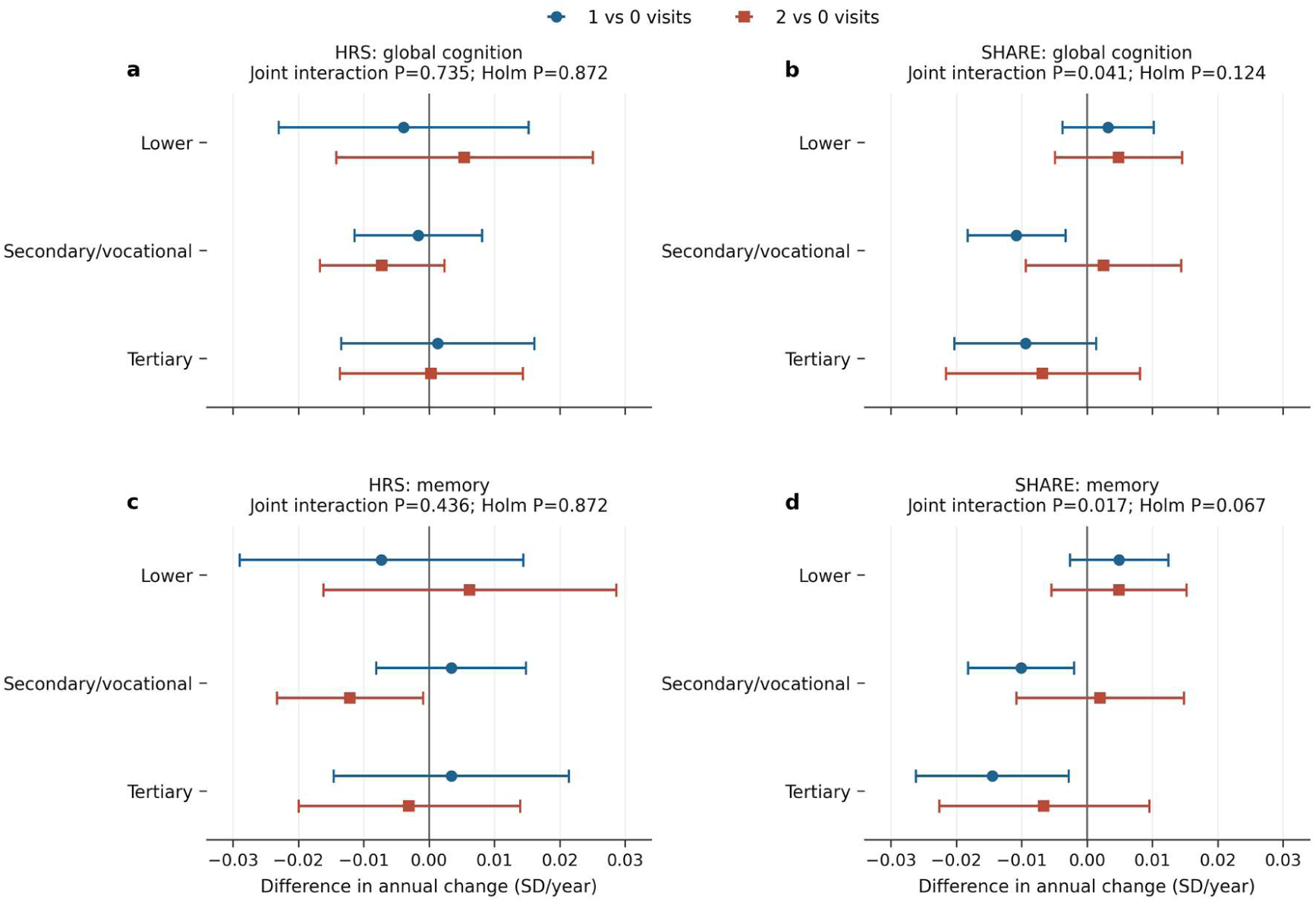
Two-wave reporting associations by educational attainment. Adjusted annual slope differences (SD/year) and nominal 95% confidence intervals within lower, upper secondary/vocational, and tertiary education strata. Omnibus four-degree-of-freedom interaction P values and familywise Holm-adjusted P values across the four education interaction tests are shown above each panel.

### Sensitivity analyses and model diagnostics

Across all supplementary sensitivity specifications, primary conclusions remained unchanged (Tables S2a, S2b, S4, S4b). Compared with participants retained in the longitudinal landmark sample, eligible individuals excluded due to missing post-landmark cognitive follow-up were older, had greater medical comorbidity and functional impairment, and exhibited lower landmark cognitive scores (ASMD = 0.519 in HRS; 0.437 in SHARE; Table S2a, Part 1). Refitting the landmark models with stabilized inverse-probability-of-inclusion weights yielded point estimates nearly identical to unweighted analyses, with all eight weighted 95% confidence intervals crossing zero (all Holm P = 0.782; Table S2a, Parts 2–3). In multivariable multinomial reverse-causation models, lower visit-1 cognition and steeper visit-1-to-landmark cognitive decline were modestly associated with subsequent single-visit reporting in selected models (HRS memory visit-1 score: OR = 0.929 per 1 SD higher score, P = 0.030; HRS memory pre-landmark change: OR = 0.935, P = 0.028; SHARE global cognition visit-1 score: OR = 0.938, P = 0.040), whereas pre-landmark cognitive change did not predict sustained two-visit reporting in any model (all P ≥ 0.124), and the smallest P for visit-1 baseline cognition was 0.067 for HRS global cognition (Table S2b). Adjusting for concurrent non-sleep, non-psychotropic medication count altered slope coefficients by <0.001 SD/year across all contrasts, and partitioning the SHARE EURO-D scale into the sleep-trouble item (euro5) and the remaining 11-item non-sleep depressive score left estimates virtually unchanged (Table S4). Finally, in negative-control analyses of antihypertensive and lipid-lowering medications, none of the 16 count-based contrasts reached nominal or familywise significance (smallest nominal P = 0.067; all Holm P = 1.000; Table S4b).

## Discussion

### Principal findings

In this harmonized prospective investigation of two large population-based aging cohorts in the United States and Europe, single-wave baseline sleep-medication reporting was associated with modestly faster subsequent decline in global cognition and memory. When exposure was evaluated across two consecutive biennial waves using an explicit landmark design, however, reporting sleep-medication use at both visits showed no association with accelerated cognitive decline in either HRS or SHARE, refuting a simple monotonic cumulative-exposure gradient. Instead, steeper post-landmark cognitive decline localized to participants reporting sleep-medication use at exactly one of the two exposure visits, concentrated in Western Europe and among SHARE participants with upper secondary/vocational or tertiary education. In the three-wave extension, the small negative HRS estimates clustered around transient patterns rather than sustained use, and equivalence testing placed sustained three-visit reporting within a ±0.05 SD/year band of non-use in both cohorts, arguing against a cumulative dose-dependent effect across a three-wave (4- to 5-year) exposure window.

### Comparison with prior literature and clinical magnitude

Our baseline estimates align in direction with prior observational studies and meta-analyses linking sedative-hypnotic prescribing to accelerated cognitive decline or dementia incidence [7–10], while clarifying the modest absolute magnitude of self-reported associations in community-dwelling populations. Across primary and subgroup analyses, annual slope differences ranged from −0.0044 to −0.0145 SD/year—substantially below the commonly referenced minimal clinically important difference (MCID) of 0.5 SD for standardized cognitive batteries. At a constant linear rate, accruing a 0.5 SD decrement would require approximately 34.5 years at the steepest observed subgroup slope (−0.0145 SD/year in SHARE tertiary education), 57.5 years at the Western European one-visit slope (−0.0087 SD/year), and more than a century at the pooled baseline slope (−0.0044 SD/year). These modest effect sizes parallel recent population-based neuroimaging and cohort studies demonstrating that while baseline hypnotic indicators often correlate with health vulnerability, cumulative dose-response relationships with neurodegeneration or long-term cognitive decline are frequently attenuated or absent after accounting for prodromal symptoms and indication [15,16]. Similar small annual slope differences have been documented for longitudinal adherence trajectories of cardiovascular medications in aging cohorts [33].

### Mechanisms underlying the single-visit transition signal

Three complementary mechanisms help explain why negative cognitive slope differences localized to intermittent single-visit reporting rather than sustained two-visit reporting. First, a depletion-of-susceptibles (or healthy-tolerator) process may operate across repeated waves: older adults who tolerate chronic sleep medication without acute cognitive or functional adverse effects are selectively retained in the two-visit group, whereas individuals experiencing emerging cognitive vulnerability, mood instability, or medication intolerance are more likely to initiate or discontinue treatment between waves. Second, intermittent sleep-medication reporting may serve as a clinical marker of prodromal sleep-wake and neuropsychiatric instability. Preclinical neurodegenerative processes frequently manifest with sleep fragmentation and affective fluctuations that prompt episodic help-seeking and short-term hypnotic prescribing; our reverse-causation finding that lower initial cognition and steeper pre-landmark cognitive decline modestly predicted single-visit (but not two-visit) reporting supports this prodromal pathway. Third, in sequence-resolved models, both initiation (01) and discontinuation (10) in SHARE exhibited similar negative point estimates relative to non-use (00), indicating that treatment transition in either direction—rather than cumulative pharmacological exposure—captures the period of heightened cognitive vulnerability. That the mid-window single-episode pattern, rather than sustained use, carried the largest HRS estimate is consistent with a transition-related process rather than cumulative neurotoxicity, a reading supported by the reverse-causation models, in which lower pre-landmark cognition predicted early discontinuation and mid-window episodes but not sustained use.

The null findings for two-visit reporting and the statistical equivalence of sustained three-visit reporting (pattern 111) to persistent non-use (within ±0.05 SD/year) across both cohorts further indicate that routine biennial self-reports of sleep-medication use do not capture a pronounced cumulative neurotoxic trajectory at the population level. Future studies integrating detailed dispensing records, daily dose equivalents, and receptor-specific drug classifications— such as the HRS prescription drug study sub-modules or linked national prescription registries in Europe—will be valuable to test whether specific high-potency benzodiazepines or strong anticholinergic hypnotics carry cumulative risks that are diluted in broad self-reported indicators.

### Educational gradient in SHARE

Within SHARE, steeper one-visit cognitive decline was observed among participants with upper secondary/vocational and tertiary education compared with those in the lower educational stratum. This pattern is consistent with two established epidemiological mechanisms. First, under the cognitive reserve hypothesis [34], individuals with higher educational attainment maintain higher baseline cognitive performance during early neuropathological accumulation but experience steeper observable decline once compensatory reserve thresholds are exceeded; conversely, lower baseline scores in less-educated strata can compress measurable longitudinal change due to floor effects, a phenomenon documented in recent multicohort cognitive and functional trajectory studies in HRS and SHARE [24]. Second, health literacy, physician access, and medication-reporting accuracy typically scale with educational attainment, whereas higher rates of non-differential recall error in lower educational strata may attenuate observed slope contrasts toward the null. Because this educational gradient was not replicated in HRS and the omnibus interaction tests in SHARE reached nominal rather than familywise significance after Holm correction, these subgroup findings warrant independent replication.

### Regional concentration in Western Europe

The localization of the SHARE single-visit signal to Western Europe is consistent with long-standing regional differences in European hypnotic prescribing and regulatory policy. Historically, Western European nations documented substantially higher benzodiazepine and Z-drug utilization among older adults than Northern or Eastern European countries [5,6], followed over the past decade by concerted clinical deprescribing initiatives and restricted reimbursement guidelines. In settings with both high historical exposure and active deprescribing pressure, older adults with emerging cognitive or clinical vulnerability are more likely to undergo treatment initiation, switching, or discontinuation across consecutive survey waves. Leave-one-country-out sensitivity analyses confirmed that the Western European association was stable across all constituent countries rather than driven by a single national sample.

Taken together, our baseline (Analysis 1), two-wave landmark (Analysis 2), and three-wave landmark (Analysis 3) results provide a coherent longitudinal picture. Single-wave baseline assessments combine long-term users with individuals undergoing acute treatment transitions and include follow-up immediately after exposure ascertainment, yielding small negative average associations across both cohorts. When exposure is resolved across two or three consecutive waves and follow-up is anchored at the post-exposure landmark, sustained multi-wave reporting shows no excess decline and is statistically equivalent to non-use, demonstrating that the apparent risk is concentrated in treatment-transition states and specific sociodemographic contexts rather than chronic reporting consistency.

### Strengths and limitations

Strengths of this study include its large, harmonized cross-national sample of older adults across the United States and Europe; reconstruction of all variables from primary cohort releases under a locked statistical analysis plan; use of explicit two-wave and three-wave landmark designs that preclude immortal-time bias; formal equivalence testing of sustained multi-wave exposure; and comprehensive robustness checks encompassing inverse-probability weighting, interview-mode adjustment, bidirectional reverse-causation modeling, concurrent polypharmacy adjustment, depressive sleep-item partitioning, and negative-control medication exposures. Several limitations also merit consideration. First, sleep-medication exposure was ascertained via biennial self-reports of regular or weekly use; neither cohort’s core harmonized module captures specific active ingredients, ATC codes, daily dosages, treatment durations, or over-the-counter sleep aids, precluding drug-class-specific comparisons (e.g., benzodiazepines vs Z-drugs or sedating antidepressants). Second, although models adjusted for baseline depressive symptoms, self-rated health, psychotropic co-medication, and sleep-specific depressive items in SHARE, observational survey data cannot fully eliminate residual confounding by indication or unmeasured objective sleep architecture. Third, complete-case covariate selection and post-landmark follow-up requirements excluded participants with higher baseline morbidity and lower cognitive scores; however, inverse-probability-weighted models in both the two-wave and three-wave designs closely reproduced the unweighted estimates. Fourth, while cognitive composites were standardized within each cohort, slight differences in test batteries (Serial 7s and backward counting in HRS vs animal fluency in SHARE) mean that cohort-specific models remain primary. Fifth, in the three-wave extension (Analysis 3), interview mode in SHARE was harmonized via a participant-level indicator of telephone/remote versus face-to-face interviewing, while the HRS subsample restricted exclusively to face-to-face interviews across waves 11–15 (n = 790) contained fewer than 100 participants in exposed cells, precluding standalone CAPI-only global-cognition estimation. Finally, because the three-wave landmark was anchored at HRS wave 13 (2016) and SHARE wave 8 (2019/20) within the analyzed public releases (extending through HRS wave 15 [2020] and SHARE wave 9 [2021/22]), post-landmark follow-up in Analysis 3 was shorter than in Analysis 2, and inverse-probability weights in Analysis 3 specifically accounted for completion of the three-wave exposure window.

### Clinical and public-health implications

Although population-average slope differences were small relative to clinical thresholds for cognitive impairment, the consistent localization of steeper decline to intermittent and transient sleep-medication reporters carries practical relevance for geriatric care. In routine practice, a new initiation, single-wave episode, or recent discontinuation of sleep medication in an older adult— particularly among educated patients in settings with high hypnotic prescribing—may signal underlying sleep-wake instability or prodromal cognitive change warranting clinical evaluation and longitudinal cognitive monitoring. At the same time, the absence of accelerated decline and formal statistical equivalence among stable two- and three-visit reporters underscore that deprescribing decisions should be individualized, balancing potential medication-related risks against the well-documented cognitive and functional consequences of untreated chronic insomnia [2,3,18,19].

## Conclusions

In harmonized analyses of HRS and SHARE, baseline sleep-medication reporting was associated with small negative differences in subsequent cognitive trajectories, whereas fixed two-wave and three-wave landmark designs demonstrated no excess cognitive decline among participants reporting sustained use across consecutive visits, with three-visit reporting statistically equivalent to persistent non-use within ±0.05 SD/year. Instead, steeper decline localized to intermittent single-visit and transient reporting patterns, particularly in Western Europe and higher educational strata in SHARE. Future pharmacoepidemiological studies should prioritize linked prescription and clinical indication data to disentangle acute treatment transitions and prodromal sleep disturbance from cumulative pharmacological effects.

## Data Availability

HRS and SHARE data are available to registered researchers subject to the access and terms-of-use requirements of the respective cohort repositories: https://hrsdata.isr.umich.edu/ and https://share-eric.eu/data/data-access. Individual-level data are not redistributed. The prespecified statistical analysis plan, analysis code, and aggregate outputs are provided in the supplementary materials, and source hashes of the cohort releases used (RAND HRS 2020 version 2, Harmonized HRS version D, Harmonized SHARE version F2, and SHARE release 9.0.0) are included in the reproducibility package.

https://share-eric.eu/data/data-access

## Abbreviations

ASMD: absolute standardized mean difference
ATC: Anatomical Therapeutic Chemical
BMI: body mass index
CAPI: computer-assisted personal interviewing
CES-D: Center for Epidemiologic Studies Depression scale
CI: confidence interval
df: degrees of freedom
EURO-D: European depression scale
HRS: Health and Retirement Study
IPW: inverse probability weighting
ISCED: International Standard Classification of Education
MCID: minimal clinically important difference
OR: odds ratio
RRR: relative risk ratio
SAP: statistical analysis plan
SD: standard deviation
SHARE: Survey of Health Ageing and Retirement in Europe
STROBE: Strengthening the Reporting of Observational Studies in Epidemiology
TOST: two one-sided tests.

## Declarations

### Ethics approval and consent to participate

The Health and Retirement Study (HRS) was approved by the Institutional Review Board at the University of Michigan. The Survey of Health, Ageing and Retirement in Europe (SHARE) was reviewed and approved by the Ethics Committee of the University of Mannheim (waves 1–4) and the Ethics Council of the Max Planck Society (wave 4 onwards). All participants provided written informed consent prior to enrollment. The present study was a secondary analysis of de-identified public-release data and involved no direct contact with participants; no additional ethics approval was required at the authors’ institutions.

### Consent for publication

Not applicable. No identifiable participant information is included.

### Availability of data and materials

HRS and SHARE data are available to registered researchers subject to the access and terms-of-use requirements of the respective cohort repositories: https://hrsdata.isr.umich.edu/ and https://share-eric.eu/data/data-access. Individual-level data are not redistributed. The prespecified statistical analysis plan, analysis code, and aggregate outputs are provided in the supplementary materials, and cryptographic source hashes of the cohort releases used (RAND HRS 2020 version 2, Harmonized HRS version D, Harmonized SHARE version F2, and SHARE release 9.0.0) are included in the reproducibility package.

### Competing interests

The authors declare that they have no competing interests.

### Funding

The authors received no specific funding for this work.

### Authors’ contributions

SW and DL conceived and designed the study, curated the data, conducted the statistical analyses, interpreted the findings, and drafted and critically revised the manuscript. Both authors read and approved the final manuscript. SW is the guarantor and accepts full responsibility for the integrity of the work, had full access to the data, and controlled the decision to publish.

## Acknowledgements

We thank the participants, interviewers, investigators, and data-management teams of the HRS and SHARE for making these longitudinal cohort resources available for scientific research. The HRS and SHARE harmonized data files were made available through the Gateway to Global Aging Data.

## Additional file 1: Supplementary methods and results

*Companion to: Repeated sleep-medication reporting patterns and cognitive change in HRS and SHARE: a prospective multicohort study*.

**Figure S1.**
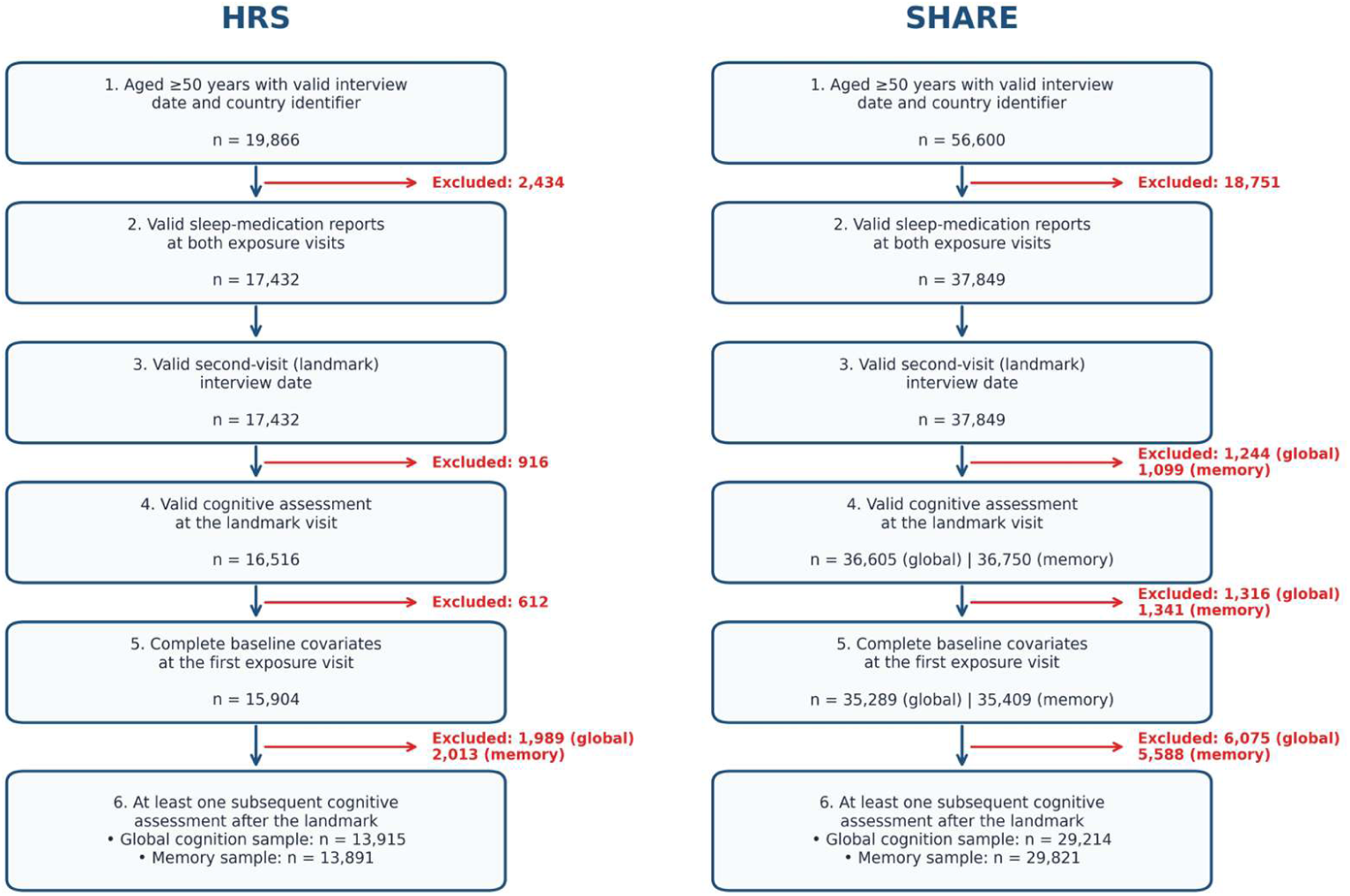
Sequential selection of the two-wave landmark analysis samples in HRS and SHARE. Six sequential inclusion criteria were applied: (1) aged ≥50 years with a valid interview date and country identifier; (2) valid sleep-medication reports at both exposure visits; (3) a valid second-visit (landmark) interview date; (4) a valid cognitive assessment at the landmark visit; (5) complete baseline covariates at the first exposure visit; and (6) at least one subsequent cognitive assessment after the landmark visit. Final analytic sample sizes are displayed for global cognition (HRS n = 13,915; SHARE n = 29,214) and memory (HRS n = 13,891; SHARE n = 29,821).

## Supplementary methods

The prespecified statistical analysis plan (SAP) is provided in the accompanying reproducibility package. Exposure classification used HRS waves 11 (2012) and 12 (2014) and SHARE waves 4 (2011) and 5 (2013), with longitudinal cognitive follow-up anchored at the second exposure visit (landmark time zero).

Full landmark models included all baseline covariates ascertained at the first exposure visit and their interactions with follow-up time; SHARE models additionally included country fixed effects and country-by-time interactions. In effect-modification models for educational attainment, the three-level education variable replaced the two baseline education indicator terms. Pooled regional interaction models in SHARE retained all four European macro-regions simultaneously. Subgroup models required at least 100 participants per exposure cell. Familywise Holm step-down adjustments were applied within each prespecified hypothesis family.

Analysis 3. The three-wave extension used three consecutive exposure visits (HRS waves 11–13; SHARE waves 6–8) and modeled cognitive trajectories from the third visit onward. Interview mode was classified as face-to-face interviewer-administered (CAPI) or remote (telephone or web in HRS; telephone in SHARE), and all models included mode and mode-by-time terms. Inverse-probability weights for completing all three exposure visits were estimated from baseline covariates, bounded at predicted probabilities of [0.02, 0.98], winsorized at the 1st and 99th percentiles, and normalized to a mean of 1.0. Equivalence of sustained use (111) to persistent non-use (000) was assessed with two one-sided tests (TOST) against margins of ±0.05 SD/year (primary) and ±0.10 SD/year (secondary). Reverse-causation models used multivariable multinomial logistic regression of the six phenotypes on pre-landmark global cognition or memory score (scaled per 1 SD higher score), fitted separately for the two cognitive domains. Corresponding results are presented in Tables S10–S13.

**Table S1.**
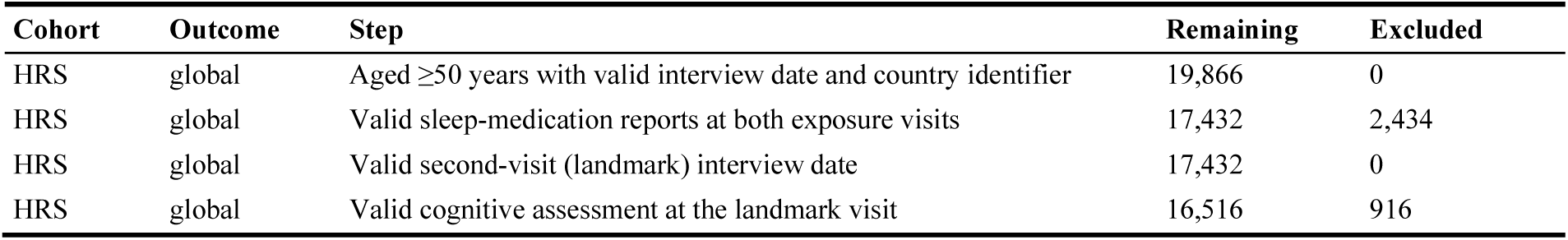
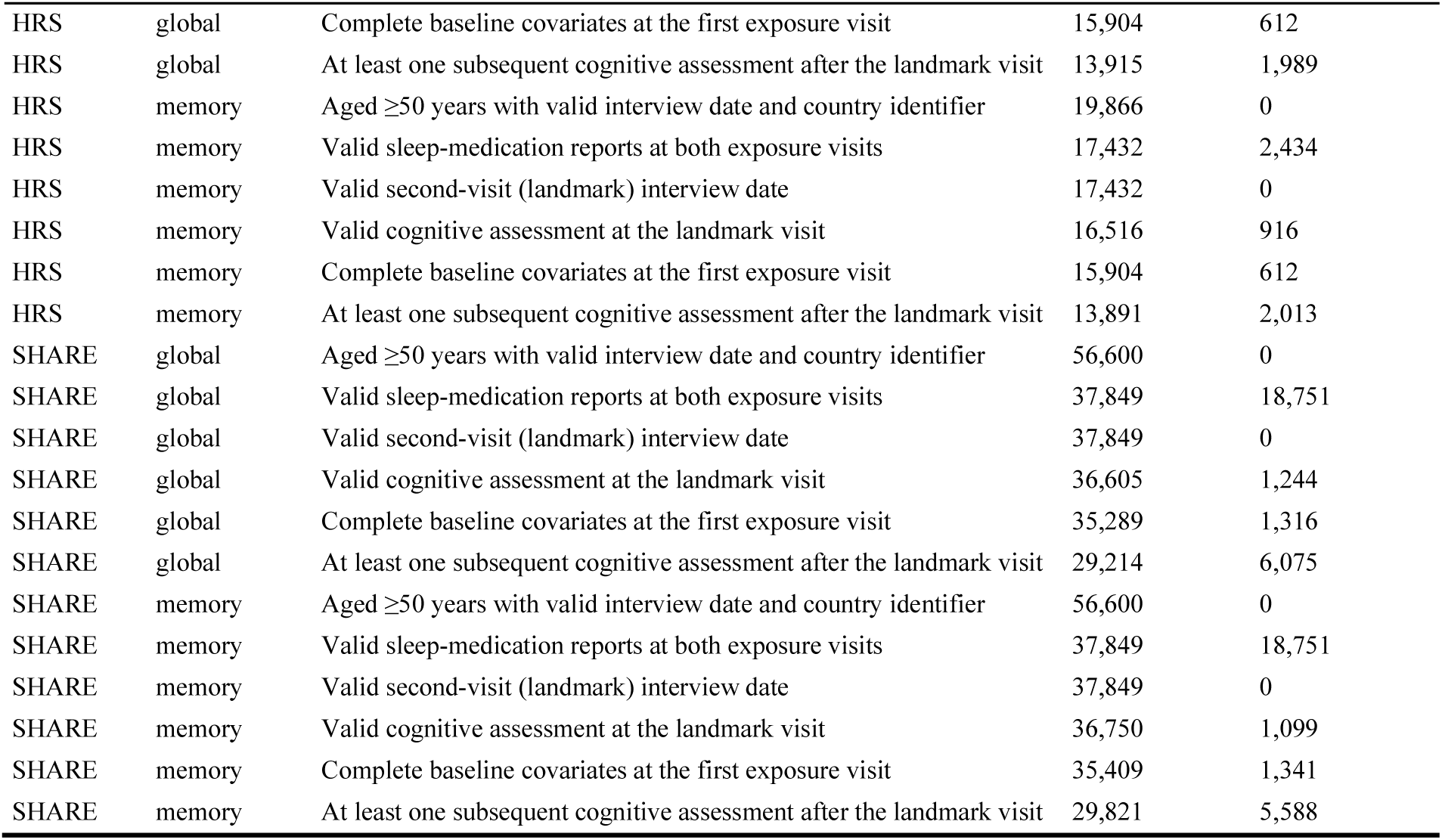
Sequential selection of two-wave landmark samples.

| Cohort | Outcome | Step | Remaining | Excluded |
|---|---|---|---|---|
| HRS | global | Aged ≥50 years with valid interview date and country identifier | 19,866 | 0 |
| HRS | global | Valid sleep-medication reports at both exposure visits | 17,432 | 2,434 |
| HRS | global | Valid second-visit (landmark) interview date | 17,432 | 0 |
| HRS | global | Valid cognitive assessment at the landmark visit | 16,516 | 916 |
| HRS | global | Complete baseline covariates at the first exposure visit | 15,904 | 612 |
| HRS | global | At least one subsequent cognitive assessment after the landmark visit | 13,915 | 1,989 |
| HRS | memory | Aged ≥50 years with valid interview date and country identifier | 19,866 | 0 |
| HRS | memory | Valid sleep-medication reports at both exposure visits | 17,432 | 2,434 |
| HRS | memory | Valid second-visit (landmark) interview date | 17,432 | 0 |
| HRS | memory | Valid cognitive assessment at the landmark visit | 16,516 | 916 |
| HRS | memory | Complete baseline covariates at the first exposure visit | 15,904 | 612 |
| HRS | memory | At least one subsequent cognitive assessment after the landmark visit | 13,891 | 2,013 |
| SHARE | global | Aged ≥50 years with valid interview date and country identifier | 56,600 | 0 |
| SHARE | global | Valid sleep-medication reports at both exposure visits | 37,849 | 18,751 |
| SHARE | global | Valid second-visit (landmark) interview date | 37,849 | 0 |
| SHARE | global | Valid cognitive assessment at the landmark visit | 36,605 | 1,244 |
| SHARE | global | Complete baseline covariates at the first exposure visit | 35,289 | 1,316 |
| SHARE | global | At least one subsequent cognitive assessment after the landmark visit | 29,214 | 6,075 |
| SHARE | memory | Aged ≥50 years with valid interview date and country identifier | 56,600 | 0 |
| SHARE | memory | Valid sleep-medication reports at both exposure visits | 37,849 | 18,751 |
| SHARE | memory | Valid second-visit (landmark) interview date | 37,849 | 0 |
| SHARE | memory | Valid cognitive assessment at the landmark visit | 36,750 | 1,099 |
| SHARE | memory | Complete baseline covariates at the first exposure visit | 35,409 | 1,341 |
| SHARE | memory | At least one subsequent cognitive assessment after the landmark visit | 29,821 | 5,588 |

**Table S2.** Two-wave exposure group and sequence counts.

| Cohort | Outcome | Exposure coding | Group | Participants | Observations |
|---|---|---|---|---|---|
| HRS | global | Number of reported-use visits (0, 1, 2) | 0 | 9,727 | 34,305 |
| HRS | global | Number of reported-use visits (0, 1, 2) | 1 | 1,970 | 6,846 |
| HRS | global | Number of reported-use visits (0, 1, 2) | 2 | 2,218 | 7,721 |
| HRS | global | Two-visit reporting sequence (00, 01, 10, 11) | 00 (No/No) | 9,727 | 34,305 |
| HRS | global | Two-visit reporting sequence (00, 01, 10, 11) | 01 (No/Yes) | 997 | 3,453 |
| HRS | global | Two-visit reporting sequence (00, 01, 10, 11) | 10 (Yes/No) | 973 | 3,393 |
| HRS | global | Two-visit reporting sequence (00, 01, 10, 11) | 11 (Yes/Yes) | 2,218 | 7,721 |
| HRS | memory | Number of reported-use visits (0, 1, 2) | 0 | 9,710 | 32,399 |
| HRS | memory | Number of reported-use visits (0, 1, 2) | 1 | 1,967 | 6,480 |
| HRS | memory | Number of reported-use visits (0, 1, 2) | 2 | 2,214 | 7,280 |
| HRS | memory | Two-visit reporting sequence (00, 01, 10, 11) | 00 (No/No) | 9,710 | 32,399 |
| HRS | memory | Two-visit reporting sequence (00, 01, 10, 11) | 01 (No/Yes) | 996 | 3,269 |
| HRS | memory | Two-visit reporting sequence (00, 01, 10, 11) | 10 (Yes/No) | 971 | 3,211 |
| HRS | memory | Two-visit reporting sequence (00, 01, 10, 11) | 11 (Yes/Yes) | 2,214 | 7,280 |
| SHARE | global | Number of reported-use visits (0, 1, 2) | 0 | 25,638 | 85,753 |
| SHARE | global | Number of reported-use visits (0, 1, 2) | 1 | 2,409 | 7,698 |
| SHARE | global | Number of reported-use visits (0, 1, 2) | 2 | 1,167 | 3,664 |
| SHARE | global | Two-visit reporting sequence (00, 01, 10, 11) | 00 (No/No) | 25,638 | 85,753 |
| SHARE | global | Two-visit reporting sequence (00, 01, 10, 11) | 01 (No/Yes) | 1,209 | 3,850 |
| SHARE | global | Two-visit reporting sequence (00, 01, 10, 11) | 10 (Yes/No) | 1,200 | 3,848 |
| SHARE | global | Two-visit reporting sequence (00, 01, 10, 11) | 11 (Yes/Yes) | 1,167 | 3,664 |
| SHARE | memory | Number of reported-use visits (0, 1, 2) | 0 | 26,139 | 99,503 |
| SHARE | memory | Number of reported-use visits (0, 1, 2) | 1 | 2,478 | 9,044 |
| SHARE | memory | Number of reported-use visits (0, 1, 2) | 2 | 1,204 | 4,297 |
| SHARE | memory | Two-visit reporting sequence (00, 01, 10, 11) | 00 (No/No) | 26,139 | 99,503 |
| SHARE | memory | Two-visit reporting sequence (00, 01, 10, 11) | 01 (No/Yes) | 1,249 | 4,532 |
| SHARE | memory | Two-visit reporting sequence (00, 01, 10, 11) | 10 (Yes/No) | 1,229 | 4,512 |
| SHARE | memory | Two-visit reporting sequence (00, 01, 10, 11) | 11 (Yes/Yes) | 1,204 | 4,297 |

**Table S2a.**
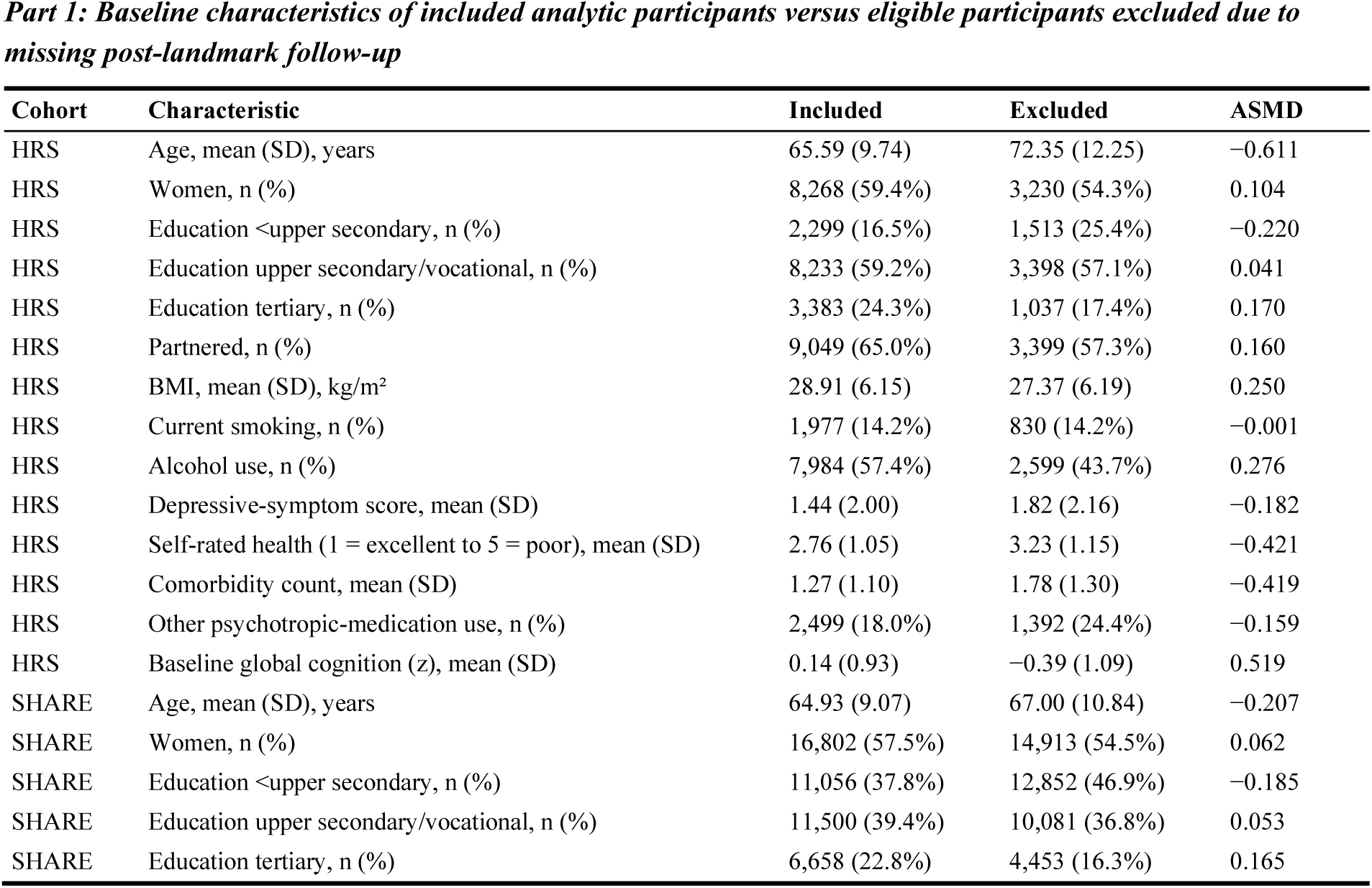
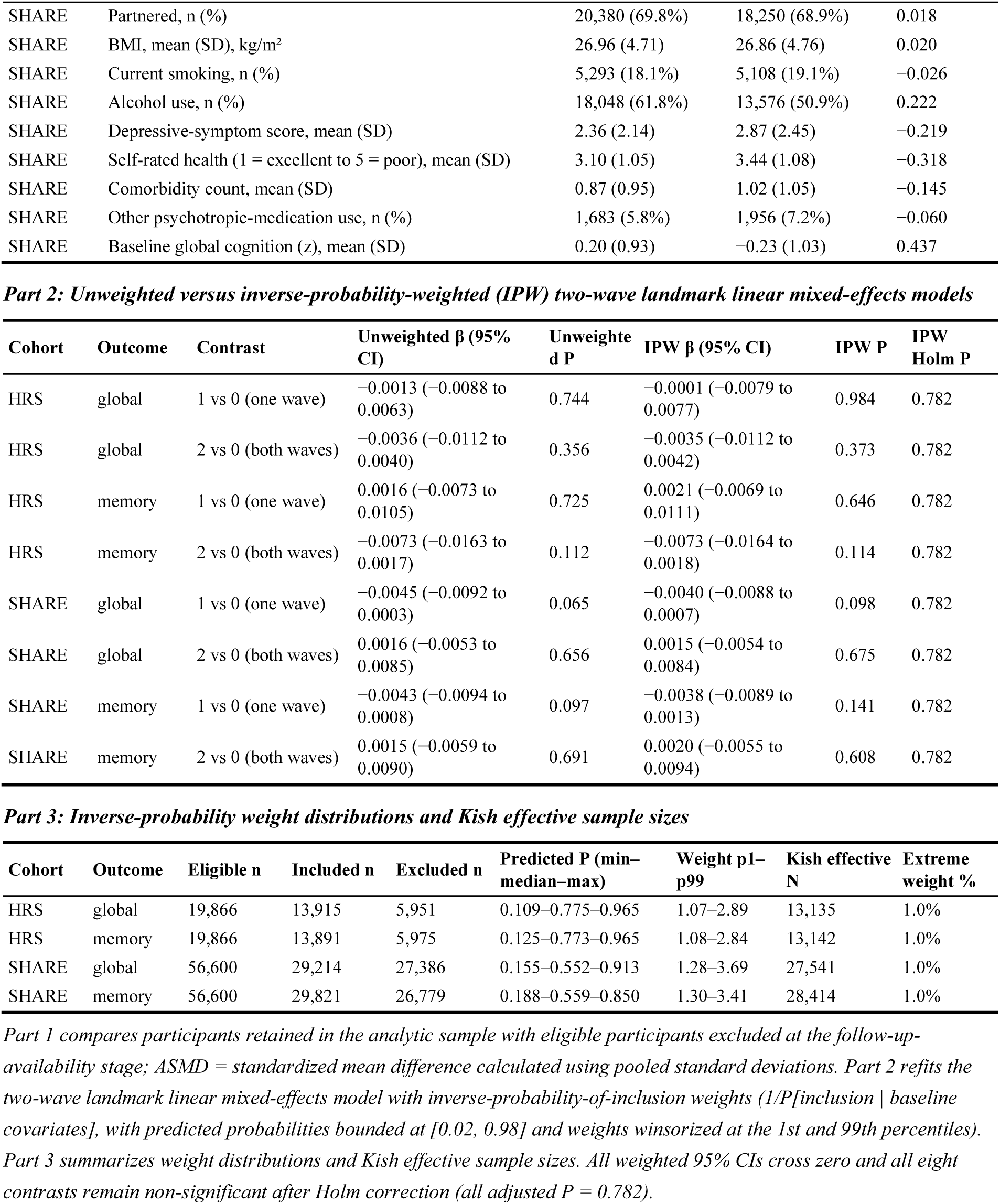
Included versus excluded baseline characteristics and inverse-probability-weighted (IPW) landmark models.

| ***Part 1: Baseline characteristics of included analytic participants versus eligible participants excluded due to missing post-landmark follow-up*** |  |  |  |  |  |  |  |  |
|---|---|---|---|---|---|---|---|---|
| **Cohort** | **Characteristic** | **Included** | **Excluded** | **ASMD** |  |  |  |  |
| HRS | Age, mean (SD), years | 65.59 (9.74) | 72.35 (12.25) | −0.611 |  |  |  |  |
| HRS | Women, n (%) | 8,268 (59.4%) | 3,230 (54.3%) | 0.104 |  |  |  |  |
| HRS | Education <upper secondary, n (%) | 2,299 (16.5%) | 1,513 (25.4%) | −0.220 |  |  |  |  |
| HRS | Education upper secondary/vocational, n (%) | 8,233 (59.2%) | 3,398 (57.1%) | 0.041 |  |  |  |  |
| HRS | Education tertiary, n (%) | 3,383 (24.3%) | 1,037 (17.4%) | 0.170 |  |  |  |  |
| HRS | Partnered, n (%) | 9,049 (65.0%) | 3,399 (57.3%) | 0.160 |  |  |  |  |
| HRS | BMI, mean (SD), kg/m² | 28.91 (6.15) | 27.37 (6.19) | 0.250 |  |  |  |  |
| HRS | Current smoking, n (%) | 1,977 (14.2%) | 830 (14.2%) | −0.001 |  |  |  |  |
| HRS | Alcohol use, n (%) | 7,984 (57.4%) | 2,599 (43.7%) | 0.276 |  |  |  |  |
| HRS | Depressive-symptom score, mean (SD) | 1.44 (2.00) | 1.82 (2.16) | −0.182 |  |  |  |  |
| HRS | Self-rated health (1 = excellent to 5 = poor), mean (SD) | 2.76 (1.05) | 3.23 (1.15) | −0.421 |  |  |  |  |
| HRS | Comorbidity count, mean (SD) | 1.27 (1.10) | 1.78 (1.30) | −0.419 |  |  |  |  |
| HRS | Other psychotropic-medication use, n (%) | 2,499 (18.0%) | 1,392 (24.4%) | −0.159 |  |  |  |  |
| HRS | Baseline global cognition (z), mean (SD) | 0.14 (0.93) | −0.39 (1.09) | 0.519 |  |  |  |  |
| SHARE | Age, mean (SD), years | 64.93 (9.07) | 67.00 (10.84) | −0.207 |  |  |  |  |
| SHARE | Women, n (%) | 16,802 (57.5%) | 14,913 (54.5%) | 0.062 |  |  |  |  |
| SHARE | Education <upper secondary, n (%) | 11,056 (37.8%) | 12,852 (46.9%) | −0.185 |  |  |  |  |
| SHARE | Education upper secondary/vocational, n (%) | 11,500 (39.4%) | 10,081 (36.8%) | 0.053 |  |  |  |  |
| SHARE | Education tertiary, n (%) | 6,658 (22.8%) | 4,453 (16.3%) | 0.165 |  |  |  |  |
| SHARE | Partnered, n (%) | 20,380 (69.8%) | 18,250 (68.9%) | 0.018 |  |  |  |  |
| SHARE | BMI, mean (SD), kg/m² | 26.96 (4.71) | 26.86 (4.76) | 0.020 |  |  |  |  |
| SHARE | Current smoking, n (%) | 5,293 (18.1%) | 5,108 (19.1%) | −0.026 |  |  |  |  |
| SHARE | Alcohol use, n (%) | 18,048 (61.8%) | 13,576 (50.9%) | 0.222 |  |  |  |  |
| SHARE | Depressive-symptom score, mean (SD) | 2.36 (2.14) | 2.87 (2.45) | −0.219 |  |  |  |  |
| SHARE | Self-rated health (1 = excellent to 5 = poor), mean (SD) | 3.10 (1.05) | 3.44 (1.08) | −0.318 |  |  |  |  |
| SHARE | Comorbidity count, mean (SD) | 0.87 (0.95) | 1.02 (1.05) | −0.145 |  |  |  |  |
| SHARE | Other psychotropic-medication use, n (%) | 1,683 (5.8%) | 1,956 (7.2%) | −0.060 |  |  |  |  |
| SHARE | Baseline global cognition (z), mean (SD) | 0.20 (0.93) | −0.23 (1.03) | 0.437 |  |  |  |  |
| ***Part 2: Unweighted versus inverse-probability-weighted (IPW) two-wave landmark linear mixed-effects models*** |  |  |  |  |  |  |  |  |
| **Cohort** | **Outcome** | **Contrast** | **Unweighted β (95% CI)** | **Unweighted P** | **IPW β (95% CI)** | **IPW P** | **IPW Holm P** |  |
| HRS | global | 1 vs 0 (one wave) | −0.0013 (−0.0088 to 0.0063) | 0.744 | −0.0001 (−0.0079 to 0.0077) | 0.984 | 0.782 |  |
| HRS | global | 2 vs 0 (both waves) | −0.0036 (−0.0112 to 0.0040) | 0.356 | −0.0035 (−0.0112 to 0.0042) | 0.373 | 0.782 |  |
| HRS | memory | 1 vs 0 (one wave) | 0.0016 (−0.0073 to 0.0105) | 0.725 | 0.0021 (−0.0069 to 0.0111) | 0.646 | 0.782 |  |
| HRS | memory | 2 vs 0 (both waves) | −0.0073 (−0.0163 to 0.0017) | 0.112 | −0.0073 (−0.0164 to 0.0018) | 0.114 | 0.782 |  |
| SHARE | global | 1 vs 0 (one wave) | −0.0045 (−0.0092 to 0.0003) | 0.065 | −0.0040 (−0.0088 to 0.0007) | 0.098 | 0.782 |  |
| SHARE | global | 2 vs 0 (both waves) | 0.0016 (−0.0053 to 0.0085) | 0.656 | 0.0015 (−0.0054 to 0.0084) | 0.675 | 0.782 |  |
| SHARE | memory | 1 vs 0 (one wave) | −0.0043 (−0.0094 to 0.0008) | 0.097 | −0.0038 (−0.0089 to 0.0013) | 0.141 | 0.782 |  |
| SHARE | memory | 2 vs 0 (both waves) | 0.0015 (−0.0059 to 0.0090) | 0.691 | 0.0020 (−0.0055 to 0.0094) | 0.608 | 0.782 |  |
| ***Part 3: Inverse-probability weight distributions and Kish effective sample sizes*** |  |  |  |  |  |  |  |  |
| **Cohort** | **Outcome** | **Eligible n** | **Included n** | **Excluded n** | **Predicted P (min–median–max)** | **Weight p1–p99** | **Kish effective N** | **Extreme weight %** |
| HRS | global | 19,866 | 13,915 | 5,951 | 0.109–0.775–0.965 | 1.07–2.89 | 13,135 | 1.0% |
| HRS | memory | 19,866 | 13,891 | 5,975 | 0.125–0.773–0.965 | 1.08–2.84 | 13,142 | 1.0% |
| SHARE | global | 56,600 | 29,214 | 27,386 | 0.155–0.552–0.913 | 1.28–3.69 | 27,541 | 1.0% |
| SHARE | memory | 56,600 | 29,821 | 26,779 | 0.188–0.559–0.850 | 1.30–3.41 | 28,414 | 1.0% |

*Part 1 compares participants retained in the analytic sample with eligible participants excluded at the follow-up-availability stage; ASMD = standardized mean difference calculated using pooled standard deviations. Part 2 refits the two-wave landmark linear mixed-effects model with inverse-probability-of-inclusion weights (1/P[inclusion \| baseline covariates], with predicted probabilities bounded at [0.02, 0.98] and weights winsorized at the 1st and 99th percentiles). Part 3 summarizes weight distributions and Kish effective sample sizes. All weighted 95% CIs cross zero and all eight contrasts remain non-significant after Holm correction (all adjusted P = 0.782).*

**Table S2b.**
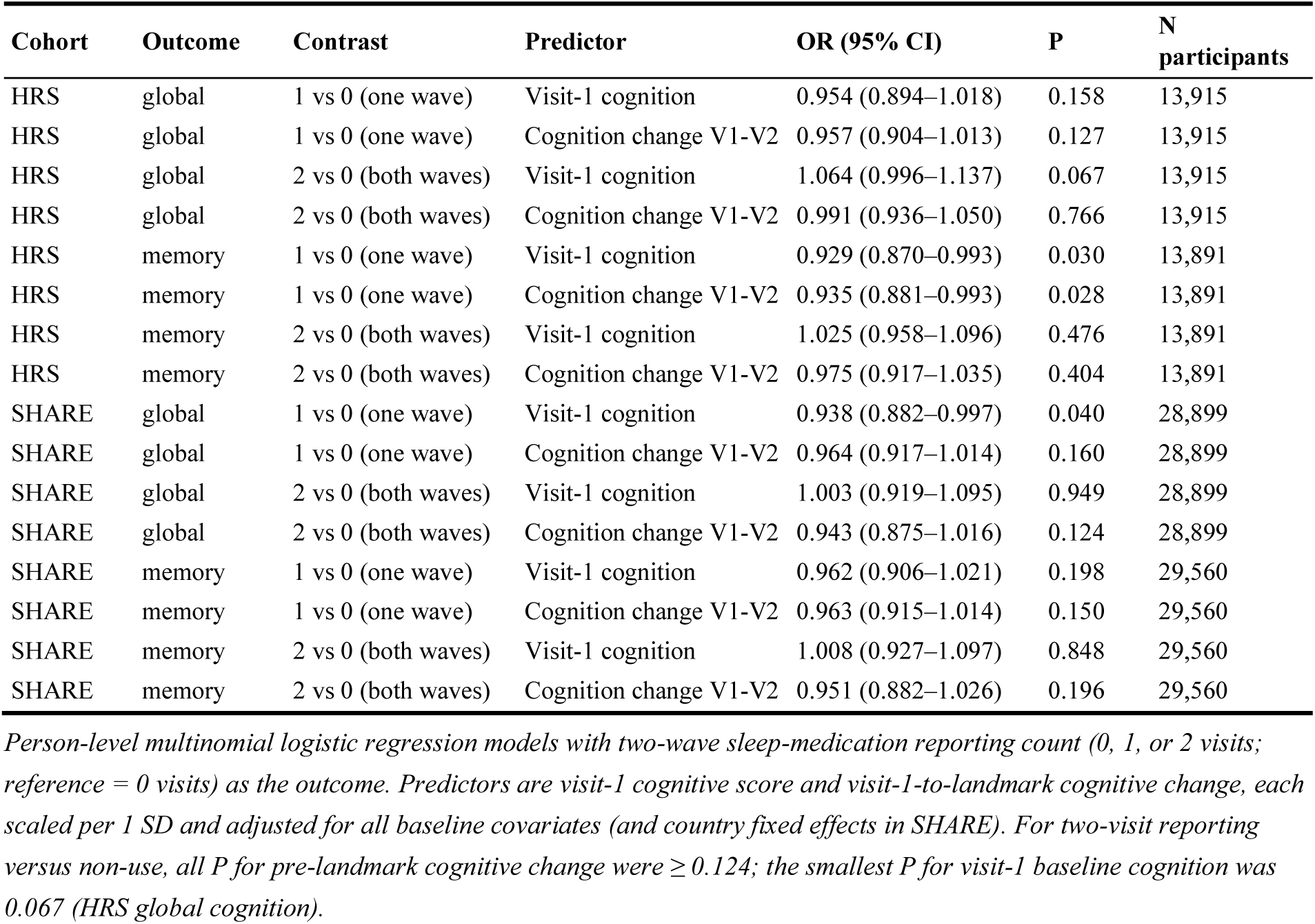
Multivariable multinomial logistic regression models evaluating reverse causation.

| Cohort | Outcome | Contrast | Predictor | OR (95% CI) | P | N participants |
|---|---|---|---|---|---|---|
| HRS | global | 1 vs 0 (one wave) | Visit-1 cognition | 0.954 (0.894–1.018) | 0.158 | 13,915 |
| HRS | global | 1 vs 0 (one wave) | Cognition change V1-V2 | 0.957 (0.904–1.013) | 0.127 | 13,915 |
| HRS | global | 2 vs 0 (both waves) | Visit-1 cognition | 1.064 (0.996–1.137) | 0.067 | 13,915 |
| HRS | global | 2 vs 0 (both waves) | Cognition change V1-V2 | 0.991 (0.936–1.050) | 0.766 | 13,915 |
| HRS | memory | 1 vs 0 (one wave) | Visit-1 cognition | 0.929 (0.870–0.993) | 0.030 | 13,891 |
| HRS | memory | 1 vs 0 (one wave) | Cognition change V1-V2 | 0.935 (0.881–0.993) | 0.028 | 13,891 |
| HRS | memory | 2 vs 0 (both waves) | Visit-1 cognition | 1.025 (0.958–1.096) | 0.476 | 13,891 |
| HRS | memory | 2 vs 0 (both waves) | Cognition change V1-V2 | 0.975 (0.917–1.035) | 0.404 | 13,891 |
| SHARE | global | 1 vs 0 (one wave) | Visit-1 cognition | 0.938 (0.882–0.997) | 0.040 | 28,899 |
| SHARE | global | 1 vs 0 (one wave) | Cognition change V1-V2 | 0.964 (0.917–1.014) | 0.160 | 28,899 |
| SHARE | global | 2 vs 0 (both waves) | Visit-1 cognition | 1.003 (0.919–1.095) | 0.949 | 28,899 |
| SHARE | global | 2 vs 0 (both waves) | Cognition change V1-V2 | 0.943 (0.875–1.016) | 0.124 | 28,899 |
| SHARE | memory | 1 vs 0 (one wave) | Visit-1 cognition | 0.962 (0.906–1.021) | 0.198 | 29,560 |
| SHARE | memory | 1 vs 0 (one wave) | Cognition change V1-V2 | 0.963 (0.915–1.014) | 0.150 | 29,560 |
| SHARE | memory | 2 vs 0 (both waves) | Visit-1 cognition | 1.008 (0.927–1.097) | 0.848 | 29,560 |
| SHARE | memory | 2 vs 0 (both waves) | Cognition change V1-V2 | 0.951 (0.882–1.026) | 0.196 | 29,560 |

*Person-level multinomial logistic regression models with two-wave sleep-medication reporting count (0, 1, or 2 visits; reference = 0 visits) as the outcome. Predictors are visit-1 cognitive score and visit-1-to-landmark cognitive change, each scaled per 1 SD and adjusted for all baseline covariates (and country fixed effects in SHARE). For two-visit reporting versus non-use, all P for pre-landmark cognitive change were ≥ 0.124; the smallest P for visit-1 baseline cognition was 0.067 (HRS global cognition).*

**Table S3.**
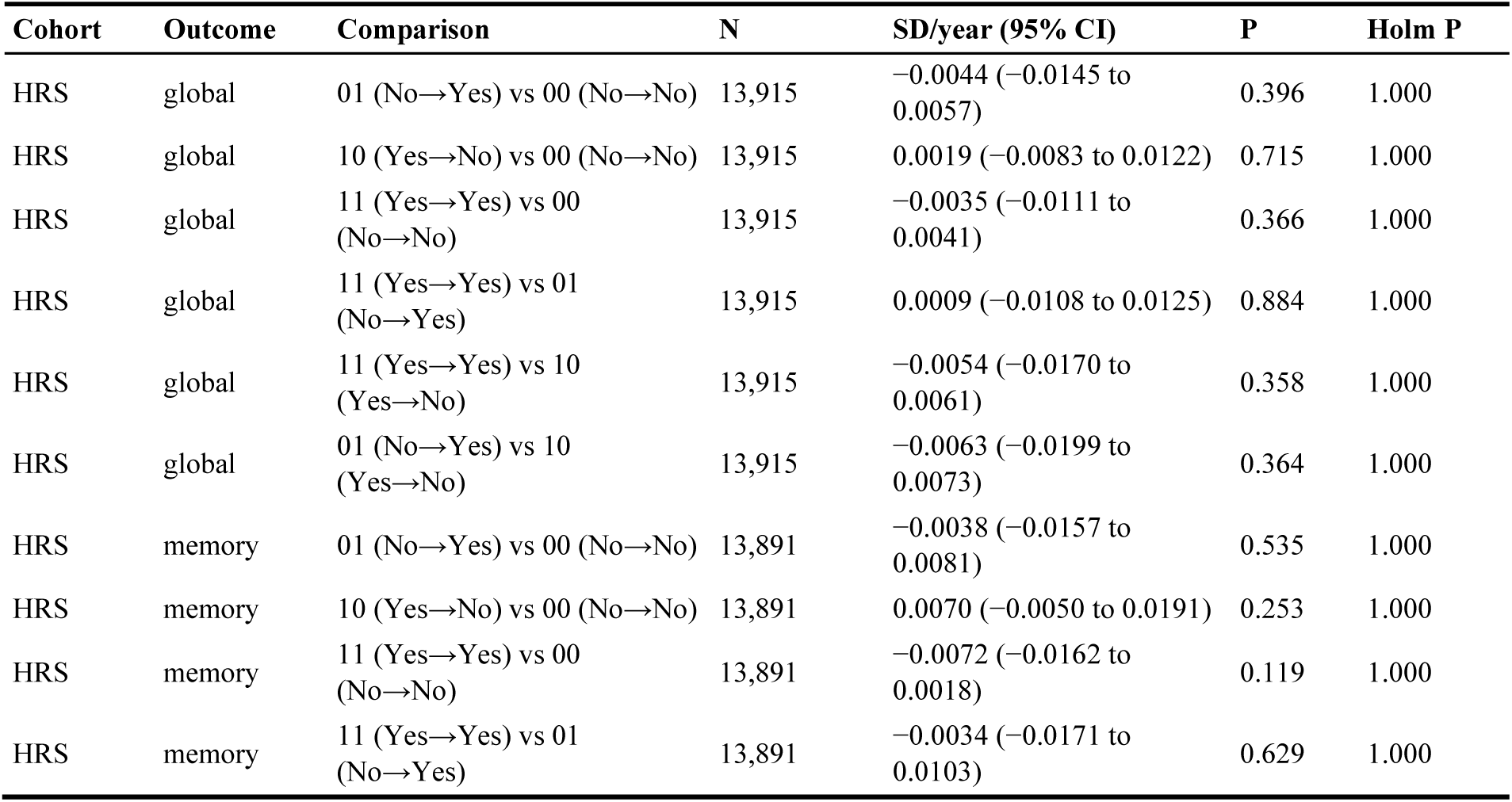
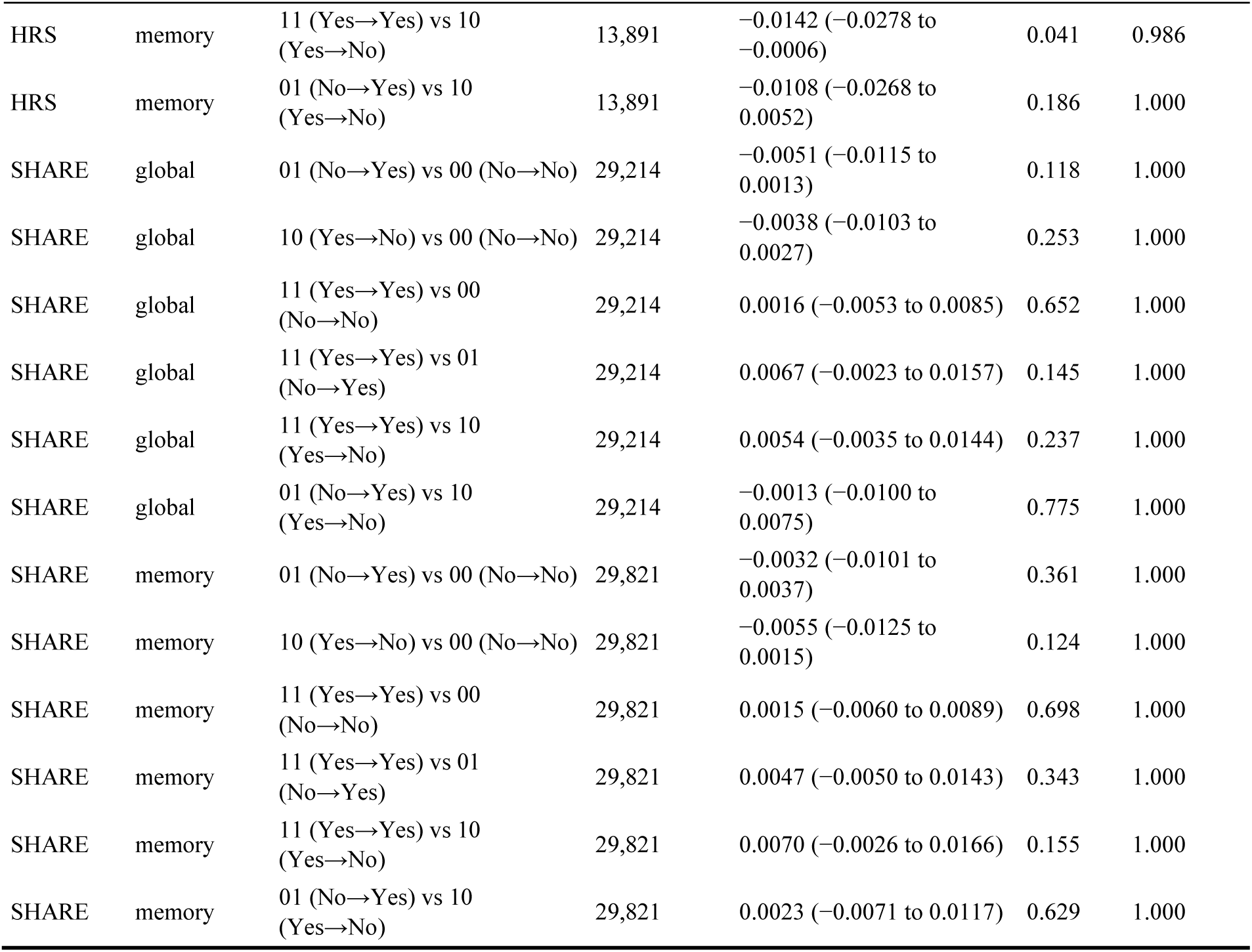
Sequence-resolved two-wave landmark estimates and direct pairwise contrasts.

**Table S4.** Core, baseline-cognition, polypharmacy-adjusted, and sleep-item split sensitivity analyses.

| Cohort | Outcome | Model specification | Comparison | N | SD/year (95% CI) | P (Holm P) |
|---|---|---|---|---|---|---|
| HRS | global | Baseline-cognition adjusted | 1 vs 0 reported-use visits | 13,915 | −0.0018 (−0.0093 to 0.0057) | 0.638 |
| HRS | global | Baseline-cognition adjusted | 2 vs 0 reported-use visits | 13,915 | −0.0042 (−0.0118 to 0.0034) | 0.279 |
| HRS | global | Core model | 1 vs 0 reported-use visits | 13,915 | −0.0020 (−0.0096 to 0.0055) | 0.595 |
| HRS | global | Core model | 2 vs 0 reported-use visits | 13,915 | −0.0054 (−0.0127 to 0.0020) | 0.153 |
| HRS | memory | Baseline-cognition adjusted | 1 vs 0 reported-use visits | 13,891 | 0.0010 (−0.0079 to 0.0099) | 0.832 |
| HRS | memory | Baseline-cognition adjusted | 2 vs 0 reported-use visits | 13,891 | −0.0083 (−0.0173 to 0.0007) | 0.070 |
| HRS | memory | Core model | 1 vs 0 reported-use visits | 13,891 | 0.0011 (−0.0077 to 0.0100) | 0.804 |
| HRS | memory | Core model | 2 vs 0 reported-use visits | 13,891 | −0.0084 (−0.0171 to 0.0003) | 0.059 |
| SHARE | global | Baseline-cognition adjusted | 1 vs 0 reported-use visits | 28,899 | −0.0040 (−0.0088 to 0.0007) | 0.097 |
| SHARE | global | Baseline-cognition adjusted | 2 vs 0 reported-use visits | 28,899 | 0.0016 (−0.0053 to 0.0085) | 0.648 |
| SHARE | global | Core model | 1 vs 0 reported-use visits | 29,214 | −0.0044 (−0.0091 to 0.0003) | 0.066 |
| SHARE | global | Core model | 2 vs 0 reported-use visits | 29,214 | 0.0017 (−0.0051 to 0.0085) | 0.629 |
| SHARE | memory | Baseline-cognition adjusted | 1 vs 0 reported-use visits | 29,560 | −0.0041 (−0.0092 to 0.0010) | 0.119 |
| SHARE | memory | Baseline-cognition adjusted | 2 vs 0 reported-use visits | 29,560 | 0.0015 (−0.0060 to 0.0089) | 0.696 |
| SHARE | memory | Core model | 1 vs 0 reported-use visits | 29,821 | −0.0041 (−0.0091 to 0.0010) | 0.115 |
| SHARE | memory | Core model | 2 vs 0 reported-use visits | 29,821 | 0.0021 (−0.0053 to 0.0094) | 0.582 |
| HRS | global | Polypharmacy-adjusted | 1 vs 0 reported-use visits | 13,915 | −0.0015 (−0.0091 to 0.0061) | 0.698 (1.000) |
| HRS | global | Polypharmacy-adjusted | 2 vs 0 reported-use visits | 13,915 | −0.0039 (−0.0116 to 0.0038) | 0.321 (1.000) |
| HRS | memory | Polypharmacy-adjusted | 1 vs 0 reported-use visits | 13,891 | 0.0013 (−0.0076 to 0.0102) | 0.774 (1.000) |
| HRS | memory | Polypharmacy-adjusted | 2 vs 0 reported-use visits | 13,891 | −0.0077 (−0.0168 to 0.0014) | 0.097 (0.615) |
| SHARE | global | Polypharmacy-adjusted | 1 vs 0 reported-use visits | 29,214 | −0.0044 (−0.0092 to 0.0003) | 0.071 (0.565) |
| SHARE | global | Polypharmacy-adjusted | 2 vs 0 reported-use visits | 29,214 | 0.0017 (−0.0052 to 0.0086) | 0.631 (1.000) |
| SHARE | memory | Polypharmacy-adjusted | 1 vs 0 reported-use visits | 29,821 | −0.0042 (−0.0093 to 0.0009) | 0.103 (0.615) |
| SHARE | memory | Polypharmacy-adjusted | 2 vs 0 reported-use visits | 29,821 | 0.0016 (−0.0058 to 0.0091) | 0.672 (1.000) |
| SHARE | global | EURO-D sleep-item split | 1 vs 0 reported-use visits | 29,101 | −0.0044 (−0.0092 to 0.0004) | 0.072 (0.231) |
| SHARE | global | EURO-D sleep-item split | 2 vs 0 reported-use visits | 29,101 | 0.0018 (−0.0051 to 0.0087) | 0.612 (1.000) |
| SHARE | memory | EURO-D sleep-item split | 1 vs 0 reported-use visits | 29,101 | −0.0046 (−0.0097 to 0.0005) | 0.058 (0.231) |
| SHARE | memory | EURO-D sleep-item split | 2 vs 0 reported-use visits | 29,101 | 0.0017 (−0.0058 to 0.0092) | 0.654 (1.000) |

*Core models omit baseline psychotropic co-medication; baseline-cognition-adjusted models additionally control for visit-1 cognitive score and its time interaction. Polypharmacy-adjusted models control for the concurrent count of non-sleep, non-psychotropic medication classes and its time interaction (HRS: 15 classes, mean 3.85 [SD 3.46]; SHARE: 10 classes, mean 2.59 [SD 2.39]). EURO-D sleep-item split models in SHARE partition the 12-item EURO-D scale into the sleep-trouble item (euro5) and the remaining 11-item non-sleep depressive score, each entered with time interactions (n = 29,101). For polypharmacy-adjusted and sleep-item split sensitivity analyses, familywise Holm-adjusted P values across the eight contrasts are reported in parentheses alongside raw P values.*

**Table S4b.** Negative-control chronic medication exposure analyses (antihypertensive and lipid-lowering medications)

| **Cohort** | **Outcome** | **Negative-control exposure** | **Visits vs 0** | **Exposed / Ref. N** | **β (SD/year)** | **95% CI** | **Nominal P** | **Holm P** |
|---|---|---|---|---|---|---|---|---|
| HRS | global | Anti-hypertensive | 1 | 3,632 / 20,520 | 0.0013 | −0.0101 to 0.0127 | 0.818 | 1.000 |
| HRS | global | Anti-hypertensive | 2 | 24,401 / 20,520 | 0.0061 | −0.0056 to 0.0179 | 0.308 | 1.000 |
| HRS | global | Lipid-lowering | 1 | 5,609 / 24,588 | 0.0080 | −0.0006 to 0.0165 | 0.067 | 1.000 |
| HRS | global | Lipid-lowering | 2 | 18,478 / 24,588 | 0.0032 | −0.0030 to 0.0094 | 0.309 | 1.000 |
| HRS | memory | Anti-hypertensive | 1 | 3,478 / 19,124 | −0.0016 | −0.0150 to 0.0117 | 0.810 | 1.000 |
| HRS | memory | Anti-hypertensive | 2 | 23,253 / 19,124 | 0.0070 | −0.0068 to 0.0208 | 0.322 | 1.000 |
| HRS | memory | Lipid-lowering | 1 | 5,360 / 23,077 | 0.0065 | −0.0035 to 0.0165 | 0.202 | 1.000 |
| HRS | memory | Lipid-lowering | 2 | 17,529 / 23,077 | 0.0029 | −0.0044 to 0.0102 | 0.435 | 1.000 |
| SHARE | global | Anti-hypertensive | 1 | 12,274 / 50,276 | 0.0037 | −0.0006 to 0.0079 | 0.088 | 1.000 |
| SHARE | global | Anti-hypertensive | 2 | 34,565 / 50,276 | −0.0004 | −0.0051 to 0.0043 | 0.868 | 1.000 |
| SHARE | global | Lipid-lowering | 1 | 12,386 / 68,821 | 0.0015 | −0.0023 to 0.0053 | 0.445 | 1.000 |
| SHARE | global | Lipid-lowering | 2 | 15,908 / 68,821 | 0.0005 | −0.0031 to 0.0041 | 0.787 | 1.000 |
| SHARE | memory | Anti-hypertensive | 1 | 14,422 / 58,348 | 0.0023 | −0.0023 to 0.0068 | 0.334 | 1.000 |
| SHARE | memory | Anti-hypertensive | 2 | 40,074 / 58,348 | −0.0007 | −0.0058 to 0.0043 | 0.772 | 1.000 |
| SHARE | memory | Lipid-lowering | 1 | 14,517 / 80,269 | 0.0002 | −0.0039 to 0.0043 | 0.921 | 1.000 |
| SHARE | memory | Lipid-lowering | 2 | 18,058 / 80,269 | 0.0007 | −0.0032 to 0.0046 | 0.727 | 1.000 |

*The identical two-wave landmark linear mixed-effects model was refit using antihypertensive and lipid-lowering medication reporting as negative-control exposures, with familywise Holm adjustment across all 16 contrasts. None of the 16 contrasts reached nominal or familywise significance (smallest nominal P = 0.067; all Holm P = 1.000). Two-visit reporting prevalence among analytic participants was 50.8% (antihypertensive) and 38.2% (lipid-lowering) in HRS, and 35.8% and 16.2% in SHARE.*

**Table S5.** Education-stratified two-wave landmark estimates.

| Cohort | Outcome | Education | Comparison | Group N | Reference N | SD/year (95% CI) | P |
|---|---|---|---|---|---|---|---|
| HRS | global | Lower | 1 vs 0 reported-use visits | 346 | 1,613 | −0.0039 (−0.0230 to 0.0152) | 0.689 |
| HRS | global | Lower | 2 vs 0 reported-use visits | 340 | 1,613 | 0.0054 (−0.0142 to 0.0250) | 0.588 |
| HRS | global | Secondary or vocational | 1 vs 0 reported-use visits | 1,175 | 5,723 | −0.0017 (−0.0114 to 0.0081) | 0.737 |
| HRS | global | Secondary or vocational | 2 vs 0 reported-use visits | 1,335 | 5,723 | −0.0072 (−0.0167 to 0.0023) | 0.140 |
| HRS | global | Tertiary | 1 vs 0 reported-use visits | 449 | 2,391 | 0.0013 (−0.0135 to 0.0161) | 0.865 |
| HRS | global | Tertiary | 2 vs 0 reported-use visits | 543 | 2,391 | 0.0003 (−0.0137 to 0.0143) | 0.964 |
| HRS | memory | Lower | 1 vs 0 reported-use visits | 346 | 1,612 | −0.0073 (−0.0290 to 0.0144) | 0.509 |
| HRS | memory | Lower | 2 vs 0 reported-use visits | 340 | 1,612 | 0.0062 (−0.0162 to 0.0286) | 0.585 |
| HRS | memory | Secondary or vocational | 1 vs 0 reported-use visits | 1,172 | 5,716 | 0.0034 (−0.0081 to 0.0148) | 0.563 |
| HRS | memory | Secondary or vocational | 2 vs 0 reported-use visits | 1,332 | 5,716 | −0.0121 (−0.0233 to −0.0009) | 0.034 |
| HRS | memory | Tertiary | 1 vs 0 reported-use visits | 449 | 2,382 | 0.0034 (−0.0146 to 0.0214) | 0.713 |
| HRS | memory | Tertiary | 2 vs 0 reported-use visits | 542 | 2,382 | −0.0031 (−0.0200 to 0.0139) | 0.723 |
| SHARE | global | Lower | 1 vs 0 reported-use visits | 1,131 | 9,307 | 0.0032 (−0.0038 to 0.0102) | 0.370 |
| SHARE | global | Lower | 2 vs 0 reported-use visits | 618 | 9,307 | 0.0048 (−0.0049 to 0.0145) | 0.332 |
| SHARE | global | Secondary or vocational | 1 vs 0 reported-use visits | 869 | 10,288 | −0.0108 (−0.0183 to −0.0033) | 0.005 |
| SHARE | global | Secondary or vocational | 2 vs 0 reported-use visits | 343 | 10,288 | 0.0025 (−0.0094 to 0.0144) | 0.682 |
| SHARE | global | Tertiary | 1 vs 0 reported-use visits | 409 | 6,043 | −0.0094 (−0.0203 to 0.0014) | 0.088 |
| SHARE | global | Tertiary | 2 vs 0 reported-use visits | 206 | 6,043 | −0.0068 (−0.0216 to 0.0081) | 0.371 |
| SHARE | memory | Lower | 1 vs 0 reported-use visits | 1,173 | 9,517 | 0.0049 (−0.0026 to 0.0124) | 0.204 |
| SHARE | memory | Lower | 2 vs 0 reported-use visits | 641 | 9,517 | 0.0049 (−0.0055 to 0.0152) | 0.358 |
| SHARE | memory | Secondary or vocational | 1 vs 0 reported-use visits | 890 | 10,492 | −0.0101 (−0.0182 to −0.0020) | 0.014 |
| SHARE | memory | Secondary or vocational | 2 vs 0 reported-use visits | 354 | 10,492 | 0.0020 (−0.0108 to 0.0148) | 0.761 |
| SHARE | memory | Tertiary | 1 vs 0 reported-use visits | 415 | 6,130 | −0.0145 (−0.0262 to −0.0028) | 0.015 |
| SHARE | memory | Tertiary | 2 vs 0 reported-use visits | 209 | 6,130 | −0.0066 (−0.0226 to 0.0095) | 0.423 |

**Table S6.** Omnibus education and regional interaction tests.

| **Cohort** | **Outcome** | **Modifier** | **Chi-square** | **df** | **P** | **Holm P** | **Family** |
|---|---|---|---|---|---|---|---|
| HRS | global | Education | 2.001 | 4 | 0.735 | 0.872 | Education (4) |
| HRS | memory | Education | 3.785 | 4 | 0.436 | 0.872 | Education (4) |
| SHARE | global | Education | 9.948 | 4 | 0.041 | 0.124 | Education (4) |
| SHARE | global | Region | 8.371 | 6 | 0.212 | 0.229 | Region (2) |
| SHARE | memory | Education | 12.066 | 4 | 0.017 | 0.067 | Education (4) |
| SHARE | memory | Region | 10.251 | 6 | 0.114 | 0.229 | Region (2) |

**Table S7.**
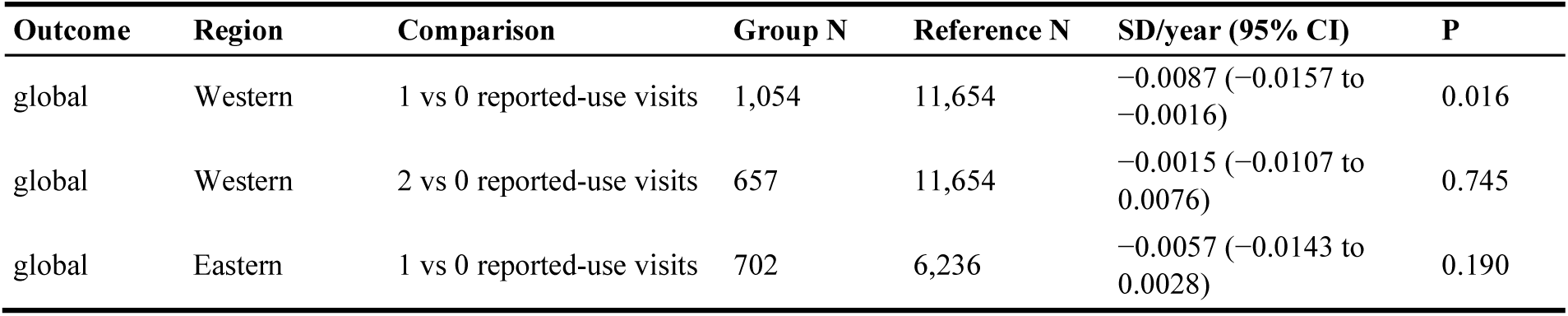
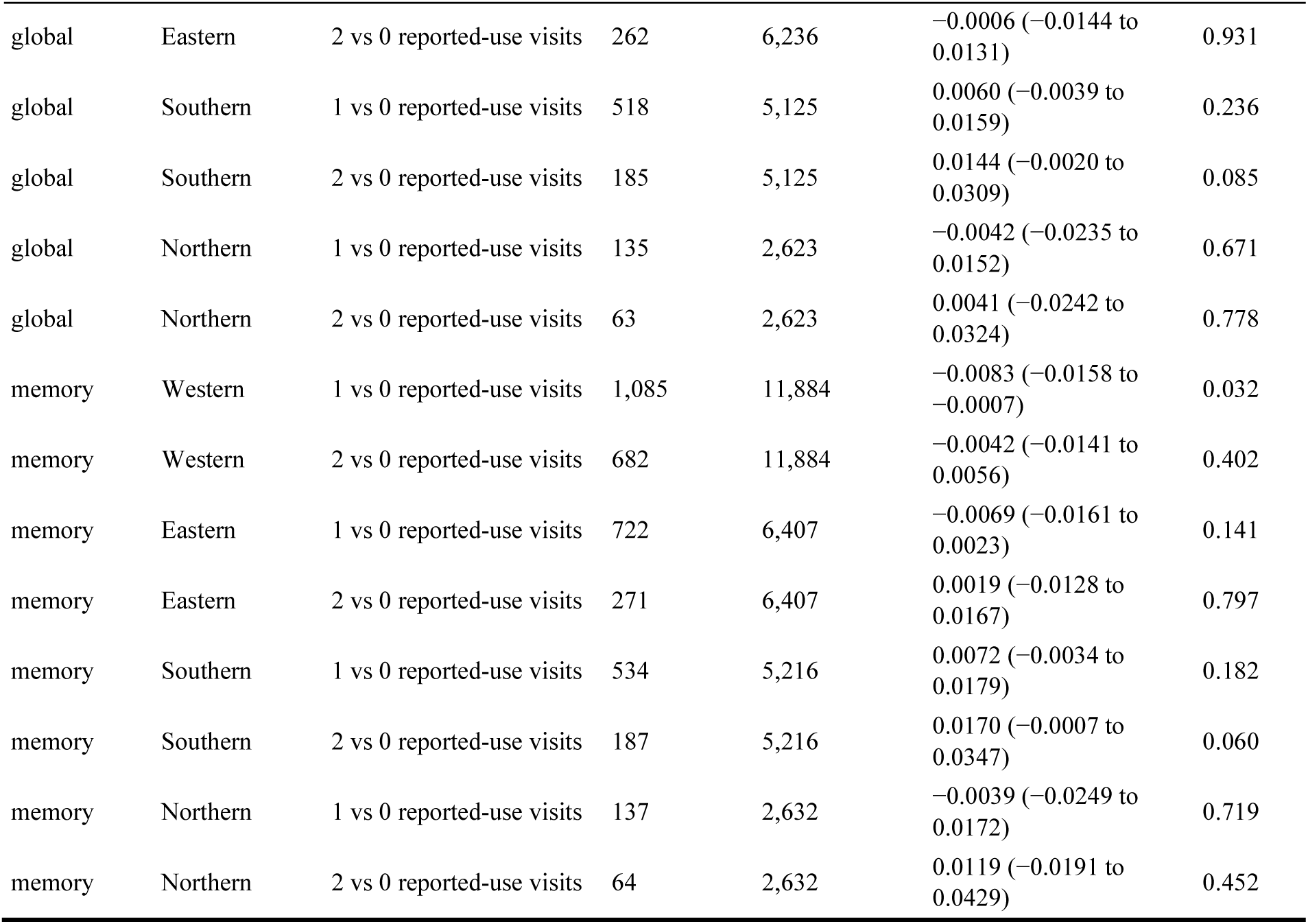
Complete SHARE regional two-wave landmark contrasts.

| Outcome | Region | Comparison | Group N | Reference N | SD/year (95% CI) | P |
|---|---|---|---|---|---|---|
| global | Western | 1 vs 0 reported-use visits | 1,054 | 11,654 | −0.0087 (−0.0157 to −0.0016) | 0.016 |
| global | Western | 2 vs 0 reported-use visits | 657 | 11,654 | −0.0015 (−0.0107 to 0.0076) | 0.745 |
| global | Eastern | 1 vs 0 reported-use visits | 702 | 6,236 | −0.0057 (−0.0143 to 0.0028) | 0.190 |
| global | Eastern | 2 vs 0 reported-use visits | 262 | 6,236 | −0.0006 (−0.0144 to 0.0131) | 0.931 |
| global | Southern | 1 vs 0 reported-use visits | 518 | 5,125 | 0.0060 (−0.0039 to 0.0159) | 0.236 |
| global | Southern | 2 vs 0 reported-use visits | 185 | 5,125 | 0.0144 (−0.0020 to 0.0309) | 0.085 |
| global | Northern | 1 vs 0 reported-use visits | 135 | 2,623 | −0.0042 (−0.0235 to 0.0152) | 0.671 |
| global | Northern | 2 vs 0 reported-use visits | 63 | 2,623 | 0.0041 (−0.0242 to 0.0324) | 0.778 |
| memory | Western | 1 vs 0 reported-use visits | 1,085 | 11,884 | −0.0083 (−0.0158 to −0.0007) | 0.032 |
| memory | Western | 2 vs 0 reported-use visits | 682 | 11,884 | −0.0042 (−0.0141 to 0.0056) | 0.402 |
| memory | Eastern | 1 vs 0 reported-use visits | 722 | 6,407 | −0.0069 (−0.0161 to 0.0023) | 0.141 |
| memory | Eastern | 2 vs 0 reported-use visits | 271 | 6,407 | 0.0019 (−0.0128 to 0.0167) | 0.797 |
| memory | Southern | 1 vs 0 reported-use visits | 534 | 5,216 | 0.0072 (−0.0034 to 0.0179) | 0.182 |
| memory | Southern | 2 vs 0 reported-use visits | 187 | 5,216 | 0.0170 (−0.0007 to 0.0347) | 0.060 |
| memory | Northern | 1 vs 0 reported-use visits | 137 | 2,632 | −0.0039 (−0.0249 to 0.0172) | 0.719 |
| memory | Northern | 2 vs 0 reported-use visits | 64 | 2,632 | 0.0119 (−0.0191 to 0.0429) | 0.452 |

**Table S8.** Western European leave-one-country-out sensitivity analyses.

| Outcome | Country omitted | Comparison | SD/year (95% CI) | P | Holm P |
|---|---|---|---|---|---|
| global | Austria | 1 vs 0 reported-use visits | −0.0098 (−0.0172 to −0.0023) | 0.011 | 0.255 |
| global | Austria | 2 vs 0 reported-use visits | −0.0005 (−0.0099 to 0.0090) | 0.921 | 1.000 |
| global | Germany | 1 vs 0 reported-use visits | −0.0078 (−0.0148 to −0.0008) | 0.029 | 0.639 |
| global | Germany | 2 vs 0 reported-use visits | −0.0005 (−0.0096 to 0.0086) | 0.918 | 1.000 |
| global | Netherlands | 1 vs 0 reported-use visits | −0.0062 (−0.0133 to 0.0009) | 0.088 | 1.000 |
| global | Netherlands | 2 vs 0 reported-use visits | −0.0017 (−0.0110 to 0.0076) | 0.717 | 1.000 |
| global | France | 1 vs 0 reported-use visits | −0.0086 (−0.0164 to −0.0008) | 0.032 | 0.662 |
| global | France | 2 vs 0 reported-use visits | −0.0033 (−0.0136 to 0.0070) | 0.527 | 1.000 |
| global | Switzerland | 1 vs 0 reported-use visits | −0.0075 (−0.0149 to −0.0000) | 0.049 | 0.879 |
| global | Switzerland | 2 vs 0 reported-use visits | −0.0012 (−0.0107 to 0.0082) | 0.800 | 1.000 |
| global | Belgium | 1 vs 0 reported-use visits | −0.0097 (−0.0179 to −0.0016) | 0.019 | 0.440 |
| global | Belgium | 2 vs 0 reported-use visits | 0.0001 (−0.0122 to 0.0124) | 0.986 | 1.000 |
| memory | Austria | 1 vs 0 reported-use visits | −0.0090 (−0.0173 to −0.0008) | 0.032 | 0.662 |
| memory | Austria | 2 vs 0 reported-use visits | −0.0027 (−0.0132 to 0.0077) | 0.605 | 1.000 |
| memory | Germany | 1 vs 0 reported-use visits | −0.0073 (−0.0149 to 0.0003) | 0.061 | 0.980 |
| memory | Germany | 2 vs 0 reported-use visits | −0.0035 (−0.0134 to 0.0063) | 0.482 | 1.000 |
| memory | Netherlands | 1 vs 0 reported-use visits | −0.0058 (−0.0135 to 0.0019) | 0.138 | 1.000 |
| memory | Netherlands | 2 vs 0 reported-use visits | −0.0049 (−0.0150 to 0.0051) | 0.338 | 1.000 |
| memory | France | 1 vs 0 reported-use visits | −0.0074 (−0.0159 to 0.0011) | 0.089 | 1.000 |
| memory | France | 2 vs 0 reported-use visits | −0.0070 (−0.0182 to 0.0042) | 0.222 | 1.000 |
| memory | Switzerland | 1 vs 0 reported-use visits | −0.0078 (−0.0159 to 0.0003) | 0.058 | 0.980 |
| memory | Switzerland | 2 vs 0 reported-use visits | −0.0046 (−0.0149 to 0.0056) | 0.373 | 1.000 |
| memory | Belgium | 1 vs 0 reported-use visits | −0.0091 (−0.0180 to −0.0003) | 0.043 | 0.818 |
| memory | Belgium | 2 vs 0 reported-use visits | 0.0003 (−0.0130 to 0.0135) | 0.969 | 1.000 |

**Table S9.** Linear mixed-effects model convergence and rank diagnostics.

| Model | Participants | Observations | Singular | Convergence | Dropped terms |
|---|---|---|---|---|---|
| HRS — Global cognition — Sequence (four-pattern) | 13,915 | 48,872 | False | OK | None |
| HRS — Global cognition — Baseline-cognition adjusted | 13,915 | 48,872 | False | OK | None |
| HRS — Global cognition — Core (without psychotropic co-medication) | 13,915 | 48,872 | False | OK | None |
| HRS — Global cognition — Education interaction | 13,915 | 48,872 | False | OK | None |
| HRS — Global cognition — Primary (count-based) | 13,915 | 48,872 | False | OK | None |
| HRS — Memory — Sequence (four-pattern) | 13,891 | 46,159 | False | OK | None |
| HRS — Memory — Baseline-cognition adjusted | 13,891 | 46,159 | False | OK | None |
| HRS — Memory — Core (without psychotropic co-medication) | 13,891 | 46,159 | False | OK | None |
| HRS — Memory — Education interaction | 13,891 | 46,159 | False | OK | None |
| HRS — Memory — Primary (count-based) | 13,891 | 46,159 | False | OK | None |
| SHARE — Global cognition — Sequence (four-pattern) | 29,214 | 97,115 | False | OK | None |
| SHARE — Global cognition — Baseline-cognition adjusted | 28,899 | 96,188 | False | OK | None |
| SHARE — Global cognition — Core (without psychotropic co-medication) | 29,214 | 97,115 | False | OK | None |
| SHARE — Global cognition — Education interaction | 29,214 | 97,115 | False | OK | None |
| SHARE — Global cognition — Primary (count-based) | 29,214 | 97,115 | False | OK | None |
| SHARE — Global cognition — Regional interaction (pooled) | 29,214 | 97,115 | False | OK | None |
| SHARE — Global cognition — Region-specific (Western) | 13,365 | 44,063 | False | OK | None |
| SHARE — Global cognition — Region-specific (Eastern) | 7,200 | 22,770 | False | OK | None |
| SHARE — Global cognition — Region-specific (Southern) | 5,828 | 19,545 | False | OK | None |
| SHARE — Global cognition — Leave-one-country-out (omitting Austria) | 10,536 | 35,478 | False | OK | None |
| SHARE — Global cognition — Leave-one-country-out (omitting Germany) | 12,444 | 40,362 | False | OK | None |
| SHARE — Global cognition — Leave-one-country-out (omitting Netherlands) | 12,119 | 40,883 | False | OK | None |
| SHARE — Global cognition — Leave-one-country-out (omitting France) | 10,457 | 34,214 | False | OK | None |
| SHARE — Global cognition — Leave-one-country-out (omitting Switzerland) | 10,968 | 35,711 | False | OK | None |
| SHARE — Global cognition — Leave-one-country-out (omitting Belgium) | 10,301 | 33,667 | False | OK | None |
| SHARE — Memory — Sequence (four-pattern) | 29,821 | 112,844 | False | OK | None |
| SHARE — Memory — Baseline-cognition adjusted | 29,560 | 111,929 | False | OK | None |
| SHARE — Memory — Core (without psychotropic co-medication) | 29,821 | 112,844 | False | OK | None |
| SHARE — Memory — Education interaction | 29,821 | 112,844 | False | OK | None |
| SHARE — Memory — Primary (count-based) | 29,821 | 112,844 | False | OK | None |
| SHARE — Memory — Regional interaction (pooled) | 29,821 | 112,844 | False | OK | None |
| SHARE — Memory — Region-specific (Western) | 13,651 | 50,667 | False | OK | None |
| SHARE — Memory — Region-specific (Eastern) | 7,400 | 28,371 | False | OK | None |
| SHARE — Memory — Region-specific (Southern) | 5,937 | 22,524 | False | OK | None |
| SHARE — Memory — Leave-one-country-out (omitting Austria) | 10,692 | 39,874 | False | OK | None |
| SHARE — Memory — Leave-one-country-out (omitting Germany) | 12,728 | 46,906 | False | OK | None |
| SHARE — Memory — Leave-one-country-out (omitting Netherlands) | 12,404 | 47,485 | False | OK | None |
| SHARE — Memory — Leave-one-country-out (omitting France) | 10,674 | 39,274 | False | OK | None |
| SHARE — Memory — Leave-one-country-out (omitting Switzerland) | 11,234 | 40,955 | False | OK | None |
| SHARE — Memory — Leave-one-country-out (omitting Belgium) | 10,523 | 38,841 | False | OK | None |

*All fitted models converged with full fixed-effect rank and non-singular random-effects covariance matrices. Standalone Northern European subgroup models were not fitted because the two-visit exposure cell contained fewer than the prespecified minimum of 100 participants; Northern Europe was retained in all pooled four-region interaction models.*

**Table S10.** Three-wave phenotype-group landmark estimates.

| **Cohort** | **Outcome** | **Phenotype group** | **N** | **SD/year (95% CI)** | **P** | **Holm P** |
|---|---|---|---|---|---|---|
| HRS | Global cognition | Late initiation | 12,310 | −0.0136 (−0.0369 to 0.0096) | 0.251 | 0.752 |
| HRS | Global cognition | Early discontinuation | 12,310 | −0.0062 (−0.0188 to 0.0064) | 0.334 | 0.752 |
| HRS | Global cognition | Mid-window single episode | 12,310 | −0.0204 (−0.0385 to −0.0023) | 0.027 | 0.137 |
| HRS | Global cognition | On-off cycling | 12,310 | 0.0175 (−0.0264 to 0.0613) | 0.435 | 0.752 |
| HRS | Global cognition | Sustained chronic use | 12,310 | −0.0146 (−0.0307 to 0.0015) | 0.076 | 0.305 |
| HRS | Memory | Late initiation | 12,310 | −0.0099 (−0.0375 to 0.0178) | 0.485 | 0.969 |
| HRS | Memory | Early discontinuation | 12,310 | −0.0099 (−0.0253 to 0.0054) | 0.205 | 0.616 |
| HRS | Memory | Mid-window single episode | 12,310 | −0.0235 (−0.0456 to −0.0013) | 0.038 | 0.189 |
| HRS | Memory | On-off cycling | 12,310 | 0.0128 (−0.0402 to 0.0659) | 0.635 | 0.969 |
| HRS | Memory | Sustained chronic use | 12,310 | −0.0183 (−0.0378 to 0.0012) | 0.066 | 0.262 |
| SHARE | Global cognition | Late initiation | 24,692 | −0.0030 (−0.0129 to 0.0068) | 0.547 | 1.000 |
| SHARE | Global cognition | Early discontinuation | 24,692 | −0.0042 (−0.0141 to 0.0057) | 0.408 | 1.000 |
| SHARE | Global cognition | Mid-window single episode | 24,692 | −0.0028 (−0.0152 to 0.0095) | 0.653 | 1.000 |
| SHARE | Global cognition | On-off cycling | 24,692 | −0.0026 (−0.0220 to 0.0168) | 0.793 | 1.000 |
| SHARE | Global cognition | Sustained chronic use | 24,692 | 0.0013 (−0.0120 to 0.0146) | 0.852 | 1.000 |
| SHARE | Memory | Late initiation | 24,692 | −0.0007 (−0.0115 to 0.0102) | 0.900 | 1.000 |
| SHARE | Memory | Early discontinuation | 24,692 | −0.0053 (−0.0162 to 0.0055) | 0.336 | 1.000 |
| SHARE | Memory | Mid-window single episode | 24,692 | 0.0024 (−0.0112 to 0.0160) | 0.731 | 1.000 |
| SHARE | Memory | On-off cycling | 24,692 | −0.0072 (−0.0286 to 0.0141) | 0.507 | 1.000 |
| SHARE | Memory | Sustained chronic use | 24,692 | −0.0006 (−0.0152 to 0.0140) | 0.936 | 1.000 |

*Exposure was classified across three consecutive visits into six phenotypes, with persistent non-use as the reference: late initiation (001 or 011), early discontinuation (100 or 110), mid-window single episode (010), on–off cycling (101), and sustained chronic use (111). SD/year values are adjusted differences in annual cognitive change from the third (landmark) visit onward; negative values indicate faster decline. Holm adjustment was applied within each cohort-outcome set across the five phenotype contrasts.*

**Table S11.**
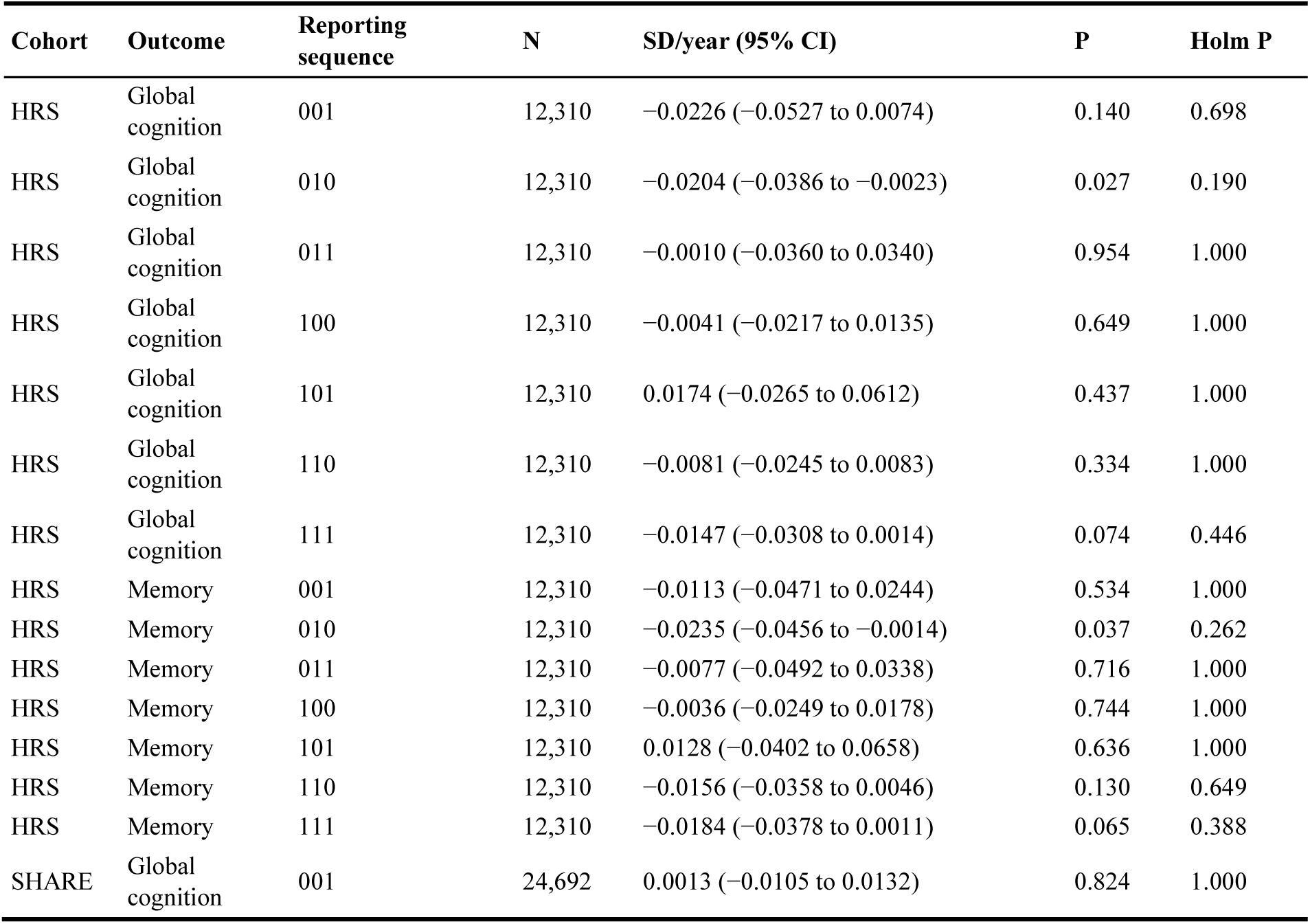
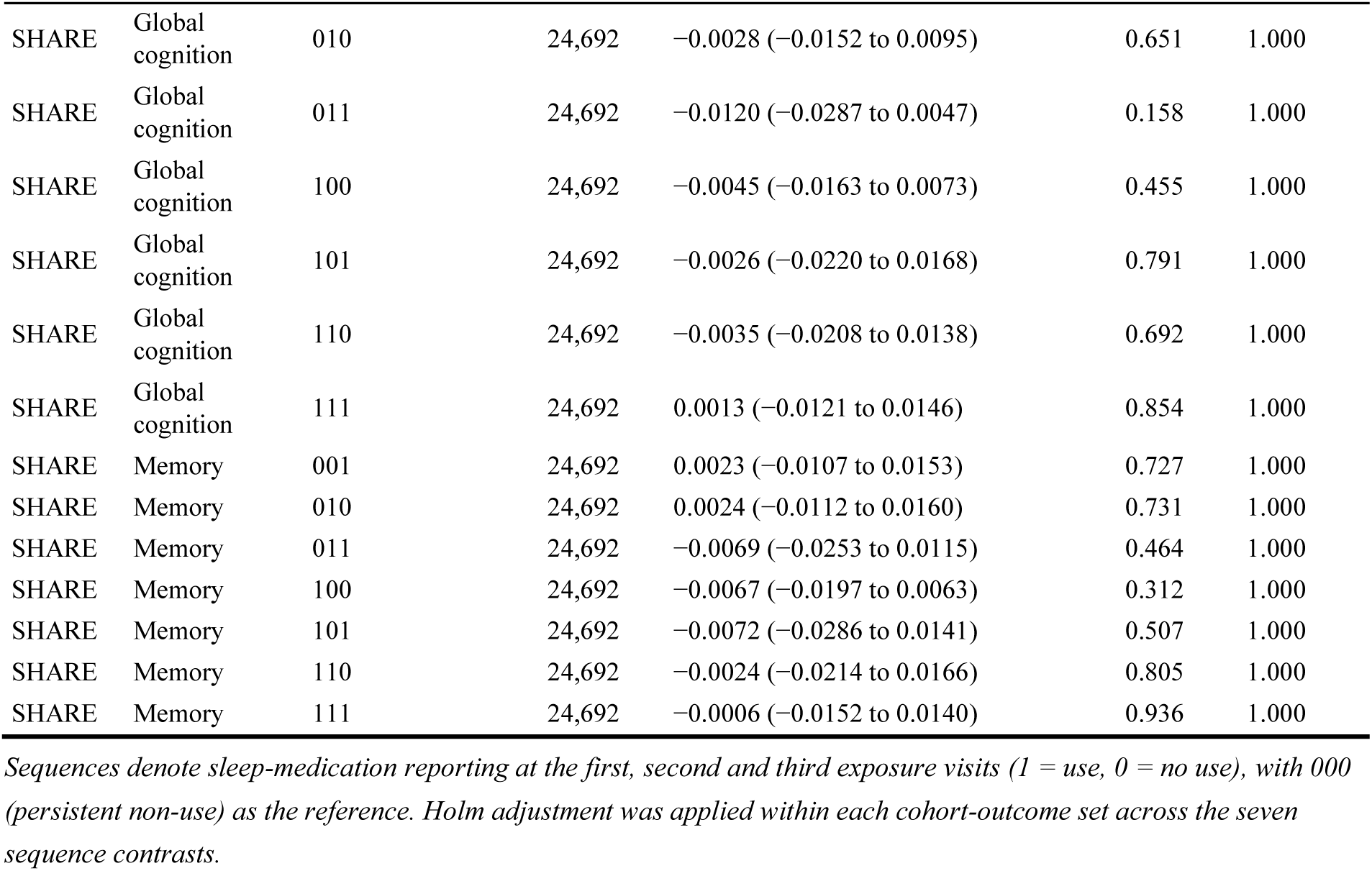
Three-wave sequence-resolved landmark estimates.

| Cohort | Outcome | Reporting sequence | N | SD/year (95% CI) | P | Holm P |
|---|---|---|---|---|---|---|
| HRS | Global cognition | 001 | 12,310 | −0.0226 (−0.0527 to 0.0074) | 0.140 | 0.698 |
| HRS | Global cognition | 010 | 12,310 | −0.0204 (−0.0386 to −0.0023) | 0.027 | 0.190 |
| HRS | Global cognition | 011 | 12,310 | −0.0010 (−0.0360 to 0.0340) | 0.954 | 1.000 |
| HRS | Global cognition | 100 | 12,310 | −0.0041 (−0.0217 to 0.0135) | 0.649 | 1.000 |
| HRS | Global cognition | 101 | 12,310 | 0.0174 (−0.0265 to 0.0612) | 0.437 | 1.000 |
| HRS | Global cognition | 110 | 12,310 | −0.0081 (−0.0245 to 0.0083) | 0.334 | 1.000 |
| HRS | Global cognition | 111 | 12,310 | −0.0147 (−0.0308 to 0.0014) | 0.074 | 0.446 |
| HRS | Memory | 001 | 12,310 | −0.0113 (−0.0471 to 0.0244) | 0.534 | 1.000 |
| HRS | Memory | 010 | 12,310 | −0.0235 (−0.0456 to −0.0014) | 0.037 | 0.262 |
| HRS | Memory | 011 | 12,310 | −0.0077 (−0.0492 to 0.0338) | 0.716 | 1.000 |
| HRS | Memory | 100 | 12,310 | −0.0036 (−0.0249 to 0.0178) | 0.744 | 1.000 |
| HRS | Memory | 101 | 12,310 | 0.0128 (−0.0402 to 0.0658) | 0.636 | 1.000 |
| HRS | Memory | 110 | 12,310 | −0.0156 (−0.0358 to 0.0046) | 0.130 | 0.649 |
| HRS | Memory | 111 | 12,310 | −0.0184 (−0.0378 to 0.0011) | 0.065 | 0.388 |
| SHARE | Global cognition | 001 | 24,692 | 0.0013 (−0.0105 to 0.0132) | 0.824 | 1.000 |
| SHARE | Global cognition | 010 | 24,692 | −0.0028 (−0.0152 to 0.0095) | 0.651 | 1.000 |
| SHARE | Global cognition | 011 | 24,692 | −0.0120 (−0.0287 to 0.0047) | 0.158 | 1.000 |
| SHARE | Global cognition | 100 | 24,692 | −0.0045 (−0.0163 to 0.0073) | 0.455 | 1.000 |
| SHARE | Global cognition | 101 | 24,692 | −0.0026 (−0.0220 to 0.0168) | 0.791 | 1.000 |
| SHARE | Global cognition | 110 | 24,692 | −0.0035 (−0.0208 to 0.0138) | 0.692 | 1.000 |
| SHARE | Global cognition | 111 | 24,692 | 0.0013 (−0.0121 to 0.0146) | 0.854 | 1.000 |
| SHARE | Memory | 001 | 24,692 | 0.0023 (−0.0107 to 0.0153) | 0.727 | 1.000 |
| SHARE | Memory | 010 | 24,692 | 0.0024 (−0.0112 to 0.0160) | 0.731 | 1.000 |
| SHARE | Memory | 011 | 24,692 | −0.0069 (−0.0253 to 0.0115) | 0.464 | 1.000 |
| SHARE | Memory | 100 | 24,692 | −0.0067 (−0.0197 to 0.0063) | 0.312 | 1.000 |
| SHARE | Memory | 101 | 24,692 | −0.0072 (−0.0286 to 0.0141) | 0.507 | 1.000 |
| SHARE | Memory | 110 | 24,692 | −0.0024 (−0.0214 to 0.0166) | 0.805 | 1.000 |
| SHARE | Memory | 111 | 24,692 | −0.0006 (−0.0152 to 0.0140) | 0.936 | 1.000 |

*Sequences denote sleep-medication reporting at the first, second and third exposure visits (1 = use, 0 = no use), with 000 (persistent non-use) as the reference. Holm adjustment was applied within each cohort-outcome set across the seven sequence contrasts.*

**Table S12.** Interview-mode restriction and inverse-probability-weighted three-wave estimates.

| Cohort | Model | Outcome | Contrast | N | SD/year (95% CI) | P |
|---|---|---|---|---|---|---|
| HRS | CAPI only | Global cognition | 1 vs 0 | 790 | Not estimated (cell n < 100) | — |
| HRS | CAPI only | Global cognition | 2 vs 0 | 790 | Not estimated (cell n < 100) | — |
| HRS | CAPI only | Global cognition | 3 vs 0 | 790 | Not estimated (cell n < 100) | — |
| HRS | CAPI only | Memory | 1 vs 0 | 790 | 0.0030 (−0.0697 to 0.0757) | 0.935 |
| HRS | CAPI only | Memory | 2 vs 0 | 790 | 0.0145 (−0.0688 to 0.0977) | 0.733 |
| HRS | CAPI only | Memory | 3 vs 0 | 790 | −0.0208 (−0.1250 to 0.0834) | 0.695 |
| HRS | IPW | Global cognition | 1 vs 0 | 12,310 | −0.0134 (−0.0258 to −0.0011) | 0.033 |
| HRS | IPW | Global cognition | 2 vs 0 | 12,310 | −0.0042 (−0.0187 to 0.0103) | 0.569 |
| HRS | IPW | Global cognition | 3 vs 0 | 12,310 | −0.0157 (−0.0315 to 0.0002) | 0.053 |
| HRS | IPW | Memory | 1 vs 0 | 12,310 | −0.0135 (−0.0284 to 0.0015) | 0.078 |
| HRS | IPW | Memory | 2 vs 0 | 12,310 | −0.0113 (−0.0290 to 0.0064) | 0.210 |
| HRS | IPW | Memory | 3 vs 0 | 12,310 | −0.0194 (−0.0385 to −0.0003) | 0.046 |
| SHARE | CAPI only | Global cognition | 1 vs 0 | 12,079 | 0.0004 (−0.0117 to 0.0124) | 0.951 |
| SHARE | CAPI only | Global cognition | 2 vs 0 | 12,079 | 0.0014 (−0.0159 to 0.0188) | 0.870 |
| SHARE | CAPI only | Global cognition | 3 vs 0 | 12,079 | −0.0069 (−0.0275 to 0.0137) | 0.513 |
| SHARE | CAPI only | Memory | 1 vs 0 | 12,079 | −0.0004 (−0.0132 to 0.0124) | 0.952 |
| SHARE | CAPI only | Memory | 2 vs 0 | 12,079 | 0.0065 (−0.0119 to 0.0249) | 0.489 |
| SHARE | CAPI only | Memory | 3 vs 0 | 12,079 | −0.0046 (−0.0264 to 0.0173) | 0.683 |
| SHARE | IPW | Global cognition | 1 vs 0 | 24,692 | −0.0018 (−0.0089 to 0.0053) | 0.614 |
| SHARE | IPW | Global cognition | 2 vs 0 | 24,692 | −0.0057 (−0.0159 to 0.0044) | 0.266 |
| SHARE | IPW | Global cognition | 3 vs 0 | 24,692 | −0.0007 (−0.0136 to 0.0121) | 0.909 |
| SHARE | IPW | Memory | 1 vs 0 | 24,692 | −0.0008 (−0.0086 to 0.0071) | 0.850 |
| SHARE | IPW | Memory | 2 vs 0 | 24,692 | −0.0040 (−0.0151 to 0.0072) | 0.482 |
| SHARE | IPW | Memory | 3 vs 0 | 24,692 | −0.0026 (−0.0167 to 0.0116) | 0.723 |

*CAPI-only models restrict the three-wave sample to face-to-face interviews; in the HRS CAPI-only subset (n = 790 across all groups), exposed subgroup cells contained fewer than the prespecified minimum of 100 participants and global-cognition mixed-effects models did not converge. IPW models apply inverse-probability-of-complete-three-wave-exposure weights (mean predicted probability 0.80 in HRS and 0.46 in SHARE; Kish effective N 12,316 and 24,018). P values are nominal and these sensitivity analyses are secondary.*

**Table S13.** Part 1 Equivalence testing of sustained three-visit use versus persistent non-use.

| Cohort | Outcome | N | SD/year (95% CI) | Equivalence P (Δ=0.05) | Equivalent at Δ=0.05 |
|---|---|---|---|---|---|
| HRS | Global cognition | 9,414 | −0.0149 (−0.0312 to 0.0014) | 1.2 × 10<sup>−5</sup> | Yes |
| HRS | Memory | 9,414 | −0.0192 (−0.0390 to 0.0006) | 0.001 | Yes |
| SHARE | Global cognition | 21,564 | 0.0021 (−0.0112 to 0.0154) | 7.9 × 10<sup>−13</sup> | Yes |
| SHARE | Memory | 21,564 | 0.0006 (−0.0141 to 0.0152) | 1.9 × 10<sup>−11</sup> | Yes |

*Two one-sided tests (TOST) against prespecified margins of ±0.05 SD/year (primary) and ±0.10 SD/year (secondary); the equivalence P value is the larger of the two one-sided P values, and all four cohort-outcome combinations also met the secondary margin (P < 0.001). One-sided non-inferiority P values were below 1 × 10<sup>−5</sup> throughout. Sustained use (111) is compared with persistent non-use (000) only, so N counts the participants in these two groups with complete covariate data and excludes the other sequences.*

**Table S13.** Part 2 Reverse-causation models: pre-landmark cognition and three-wave phenotypes.

| **Cohort** | **Phenotype group** | **N** | **Global RRR (95% CI)** | **P** | **Memory RRR (95% CI)** | **P** |
|---|---|---|---|---|---|---|
| HRS | Late initiation | 8,834 | 0.94 (0.82–1.08) | 0.408 | 0.97 (0.87–1.08) | 0.564 |
| HRS | Early discontinuation | 8,834 | 0.69 (0.59–0.80) | <0.001 | 0.81 (0.71–0.92) | 0.001 |
| HRS | Mid-window single episode | 8,834 | 0.72 (0.55–0.94) | 0.014 | 0.79 (0.63–0.99) | 0.039 |
| HRS | On-off cycling | 8,834 | 0.99 (0.88–1.12) | 0.904 | 0.98 (0.89–1.08) | 0.647 |
| HRS | Sustained chronic use | 8,834 | 0.96 (0.88–1.04) | 0.293 | 1.04 (0.98–1.11) | 0.230 |
| SHARE | Late initiation | 8,929 | 1.03 (0.76–1.42) | 0.830 | 1.06 (0.86–1.32) | 0.585 |
| SHARE | Early discontinuation | 8,929 | 0.83 (0.64–1.08) | 0.167 | 0.93 (0.78–1.11) | 0.432 |
| SHARE | Mid-window single episode | 8,929 | 1.05 (0.67–1.62) | 0.844 | 0.99 (0.74–1.34) | 0.973 |
| SHARE | On-off cycling | 8,929 | 0.91 (0.68–1.23) | 0.557 | 0.96 (0.79–1.18) | 0.725 |
| SHARE | Sustained chronic use | 8,929 | 1.22 (1.01–1.48) | 0.044 | 1.21 (1.06–1.38) | 0.004 |

*Multinomial logistic regression with persistent non-use (000) as the reference category; relative risk ratios (RRR) are per 1 SD higher pre-landmark global cognition or memory score (values < 1.00 indicate that lower pre-landmark cognitive scores are associated with higher relative risk of the corresponding exposure phenotype), adjusted for baseline age, sex, education, depressive symptoms, and chronic conditions. The two cognitive domains were modeled separately to avoid collinearity.*

**Table S14.** STROBE statement checklist for cohort studies.

| **Item No** | **Recommendation (cohort study checklist)** | **Reported in this manuscript** |
|---|---|---|
| 1a | Indicate the study’s design with a commonly used term in the title or the abstract | Title; Abstract |
| 1b | Provide in the abstract an informative and balanced summary of what was done and what was found | Abstract |
| 2 | Explain the scientific background and rationale for the investigation being reported | Background, paragraphs 1–6 |
| 3 | State specific objectives, including any prespecified hypotheses | Background, paragraph 8 |
| 4 | Present key elements of study design early in the paper | Methods, Study design and analytical framework |
| 5 | Describe the setting, locations, and relevant dates, including periods of recruitment, exposure, follow-up, and data collection | Methods, Data sources and participants |
| 6 | Give the eligibility criteria, and the sources and methods of selection of participants | Methods, Data sources and participants; Figure S1; Table S1 |
| 7 | Clearly define all outcomes, exposures, predictors, potential confounders, and effect modifiers | Methods, Exposure and landmark; Cognitive outcomes and covariates; Tables S10, S11 |
| 8 | For each variable of interest, give sources of data and details of methods of assessment; describe comparability of assessment methods | Methods, Data sources and participants; Cognitive outcomes and covariates |
| 9 | Describe any efforts to address potential sources of bias | Methods, Statistical analysis; Discussion, Strengths and limitations; Tables S2, S2a, S2b, S4, S4b, S10–S13 |
| 10 | Explain how the study size was arrived at | Methods, Data sources and participants; Figure S1 |
| 11 | Explain how quantitative variables were handled in the analyses | Methods, Cognitive outcomes and covariates; Statistical analysis |
| 12 | Describe all statistical methods, including those for subgroups and interactions, missing data, sensitivity analyses, and other analyses | Methods, Statistical analysis; Tables S2, S2a, S2b, S4, S4b, S6, S8, S10–S13 |
| 13 | Report numbers of individuals at each stage of study (e.g., numbers potentially eligible, examined for eligibility, confirmed eligible, included); give reasons for non-participation; consider use of a flow diagram | Results; Figure S1; Table S1 |
| 14 | Give characteristics of study participants and information on exposures; indicate numbers with outcome data and follow-up | Results; Table 1 |
| 15 | Report numbers of outcome events or summary measures over time | Results; Tables 2, S2–S8, S10–S13 |
| 16 | Give unadjusted and adjusted estimates and their precision; report category boundaries when continuous variables were categorised; translate estimates of relative risk into absolute risk for a meaningful time period | Results; Tables 2, S2–S8, S10–S13 |
| 17 | Report other analyses done—e.g., analyses of subgroups and interactions, and sensitivity analyses | Results; Tables S2, S2a, S2b, S4, S4b, S5–S8, S10–S13 |
| 18 | Summarise key results with reference to study objectives | Discussion, paragraph 1 |
| 19 | Discuss limitations of the study, taking into account sources of potential bias or imprecision | Discussion, Strengths and limitations |
| 20 | Give a cautious overall interpretation of results considering objectives, limitations, multiplicity of analyses, results from similar studies, and other relevant evidence | Discussion, paragraphs 2–6 |
| 21 | Discuss the generalisability (external validity) of the study results | Discussion, paragraphs 5–9 |
| 22 | Give the source of funding and the role of the funders for the present study | Declarations, Funding; Declarations, Authors' contributions |

*The checklist is adapted from the STROBE statement for observational cohort studies [32] and mapped to the sections and tables of the present manuscript.*

## Notes

### Competing Interest Statement

The authors have declared no competing interest.

### Author Declarations

The Health and Retirement Study (HRS) was approved by the Institutional Review Board at the University of Michigan. The Survey of Health, Ageing and Retirement in Europe (SHARE) was reviewed and approved by the Ethics Committee of the University of Mannheim and the Ethics Council of the Max Planck Society. All participants provided written informed consent prior to enrollment. The present study was a secondary analysis of de-identified public-release survey data and involved no direct contact with participants; no additional ethics approval was required at the authors' institutions. Data were obtained from official repositories (https://hrsdata.isr.umich.edu/ and https://share-eric.eu/data/data-access).

## References

1. GBD 2019 Dementia Forecasting Collaborators. Estimation of the global prevalence of dementia in 2019 and forecasted prevalence in 2050: an analysis for the Global Burden of Disease Study 2019. Lancet Public Health. 2022;7(2):e105–e125. 10.1016/S2468-2667(21)00249-8.

2. Endomba FT, Tchebegna PY, Chiabi E, Angong Wouna DL, Guillet C, Chauvet-Gélinier JC. Epidemiology of insomnia disorder in older persons according to the Diagnostic and Statistical Manual of Mental Disorders: a systematic review and meta-analysis. Eur Geriatr Med. 2023;14(6):1261–1272. 10.1007/s41999-023-00862-2.

3. Livingston G, Huntley J, Liu KY, Costafreda SG, Selbæk G, Alladi S, et al. Dementia prevention, intervention, and care: 2024 report of the Lancet standing Commission. Lancet. 2024;404(10452):572–628. 10.1016/S0140-6736(24)01296-0.

4. National Center for Health Statistics. Use of sleep aids among adults age 18 and older: United States, 2024. NCHS Health E-Stat 116. Hyattsville, MD: National Center for Health Statistics; 2026. https://www.cdc.gov/nchs/data/hestat/hestat116.htm. Accessed 27 October 2026.

5. Luta X, Bagnoud C, Lambiris M, Aujoulat I, Boland B, Chiolero A, et al. Patterns of benzodiazepine prescription among older adults in Switzerland: a cross-sectional analysis of claims data. BMJ Open. 2020;10(1):e031156. 10.1136/bmjopen-2019-031156.

6. Lukačišinová A, Fialová D, Peel NM, Hubbard RE, Brkić J, Onder G, et al. The prevalence and prescribing patterns of benzodiazepines and Z-drugs in older nursing home residents in different European countries and Israel: retrospective results from the EU SHELTER study. BMC Geriatr. 2021;21:277. 10.1186/s12877-021-02213-x.

7. Islam MM, Iqbal U, Walther B, Atique S, Dubey NK, Nguyen PA, et al. Benzodiazepine use and risk of dementia in the elderly population: a systematic review and meta-analysis. Neuroepidemiology. 2016;47(3–4):181–191. 10.1159/000454881.

8. He Q, Chen X, Wu T, Li L, Fei X. Risk of dementia in long-term benzodiazepine users: evidence from a meta-analysis of observational studies. J Clin Neurol. 2019;15(1):9–19. 10.3988/jcn.2019.15.1.9.

9. Billioti de Gage S, Bégaud B, Bazin F, Verdoux H, Dartigues JF, Pérès K, et al. Benzodiazepine use and risk of dementia: prospective population based study. BMJ. 2012;345:e6231. 10.1136/bmj.e6231.

10. Billioti de Gage S, Moride Y, Ducruet T, Kurth T, Verdoux H, Tournier M, et al. Benzodiazepine use and risk of Alzheimer’s disease: case-control study. BMJ. 2014;349:g5205. 10.1136/bmj.g5205.

11. Gray SL, Dublin S, Yu O, Walker R, Anderson M, Hubbard RA, et al. Benzodiazepine use and risk of incident dementia or cognitive decline: prospective population based study. BMJ. 2016;352:i90. 10.1136/bmj.i90.

12. Robbins R, DiClemente RJ, Troxel AB, Jean-Louis G, Butler M, Rapoport DM, et al. Sleep medication use and incident dementia in a nationally representative sample of older adults in the US. Sleep Med. 2021;79:183–189. 10.1016/j.sleep.2020.11.004.

13. Full KM, Pusalavidyasagar S, Palta P, Sullivan KJ, Shin JI, Gottesman RF, et al. Associations of late-life sleep medication use with incident dementia in the Atherosclerosis Risk in Communities Study. J Gerontol A Biol Sci Med Sci. 2023;78(3):438–446. 10.1093/gerona/glac088.

14. Leng Y, Stone KL, Yaffe K. Race differences in the association between sleep medication use and risk of dementia. J Alzheimers Dis. 2023;91(3):1133–1139. 10.3233/JAD-221006.

15. vom Hofe I, Stricker BH, Vernooij MW, Ikram MK, Ikram MA, Wolters FJ. Benzodiazepine use in relation to long-term dementia risk and imaging markers of neurodegeneration: a population-based study. BMC Med. 2024;22:266. 10.1186/s12916-024-03437-5.

16. Osler M, Jørgensen MB. Associations of benzodiazepines, Z-drugs, and other anxiolytics with subsequent dementia in patients with affective disorders: a nationwide cohort and nested case-control study. Am J Psychiatry. 2020;177(6):497–505. 10.1176/appi.ajp.2019.19030315.

17. Kaufmann CN, Bondi MW, Thompson WK, Spira AP, Ancoli-Israel S, Malhotra A. Cognitive performance trajectories before and after sleep treatment initiation in middle-aged and older adults: results from the Health and Retirement Study. J Gerontol A Biol Sci Med Sci. 2022;77(3):570–578. 10.1093/gerona/glab164.

18. Ma Y, Liang L, Zheng F, Shi L, Zhong B, Xie W. Association between sleep duration and cognitive decline. JAMA Netw Open. 2020;3(9):e2013573. 10.1001/jamanetworkopen.2020.13573.

19. Sabia S, Fayosse A, Dumurgier J, van Hees VT, Paquet C, Sommerlad A, et al. Association of sleep duration in middle and old age with incidence of dementia. Nat Commun. 2021;12:2289. 10.1038/s41467-021-22354-2.

20. Coupland CAC, Hill T, Dening T, Morriss R, Moore M, Hippisley-Cox J. Anticholinergic drug exposure and the risk of dementia: a nested case-control study. JAMA Intern Med. 2019;179(8):1084–1093. 10.1001/jamainternmed.2019.0677.

21. Richardson K, Fox C, Maidment I, Howard R, Loke YK, Grossi CM, et al. Anticholinergic drugs and risk of dementia: case-control study. BMJ. 2018;361:k1315. 10.1136/bmj.k1315.

22. Sun L, Zhang J, Li W, Sheng J, Xiao S. Neutrophil activation may trigger tau burden contributing to cognitive progression of chronic sleep disturbance in elderly individuals not living with dementia. BMC Med. 2023;21:205. 10.1186/s12916-023-02910-x.

23. Du M, Tao L, Liu M, Liu J. Trajectories of health conditions and their associations with the risk of cognitive impairment among older adults: insights from a national prospective cohort study. BMC Med. 2024;22:23. 10.1186/s12916-024-03245-x.

24. Lin J, Kivimäki M, Zhang Y, Zhou Y, Xu X. Functional decline in the final years of life: a multicohort study. BMC Med. 2026;24:548. 10.1186/s12916-026-05057-7.

25. Nakash M, Ojalehto Lindfors E, Zhan Y, Dahl Aslan AK, Reynolds CA, Ler P, et al. Dementia risk by metabolic health and obesity in two prospective cohorts. BMC Med. 2026;24:366. 10.1186/s12916-026-05002-8.

26. Health and Retirement Study. Public survey data. https://hrsdata.isr.umich.edu/data-products/public-survey-data. Accessed 27 October 2026.

27. SHARE-ERIC. Data documentation. https://share-eric.eu/data/data-documentation. Accessed 27 October 2026.

28. Health and Retirement Study. Imputation of Cognitive Functioning Measures 1992–2020 Data Description. https://hrs.isr.umich.edu/sites/default/files/biblio/COGIMP9220_dd.pdf. Accessed 27 October 2026.

29. SHARE-ERIC. Scales and Multi-Item Indicators in the Survey of Health, Ageing and Retirement in Europe. Release 8.0.0. https://share-eric.eu/fileadmin/user_upload/Other_Publications/ScalesManual_rel.8-0-0.pdf. Accessed 27 October 2026.

30. Sonnega A, Faul JD, Ofstedal MB, Langa KM, Phillips JWR, Weir DR. Cohort profile: the Health and Retirement Study (HRS). Int J Epidemiol. 2014;43(2):576–585. 10.1093/ije/dyu067.

31. Börsch-Supan A, Brandt M, Hunkler C, Kneip T, Korbmacher J, Malter F, et al. Data resource profile: the Survey of Health, Ageing and Retirement in Europe (SHARE). Int J Epidemiol. 2013;42(4):992–1001. 10.1093/ije/dyt088.

32. von Elm E, Altman DG, Egger M, Pocock SJ, Gøtzsche PC, Vandenbroucke JP. The Strengthening the Reporting of Observational Studies in Epidemiology (STROBE) statement: guidelines for reporting observational studies. Ann Intern Med. 2007;147(8):573–577. 10.7326/0003-4819-147-8-200710160-00010.

33. Li C, He D, Liu Y, Yang C, Zhang L. Anti-hypertensive medication adherence, socioeconomic status, and cognitive aging in the Chinese community-dwelling middle-aged and older adults ≥45 years: a population-based longitudinal study. BMC Med. 2025;23:121. 10.1186/s12916-025-03949-8.

34. Stern Y. Cognitive reserve in ageing and Alzheimer’s disease. Lancet Neurol. 2012;11(11):1006– 1012. 10.1016/S1474-4422(12)70191-6.

